# NeoScore integrates characteristics of the neoantigen:MHC class I interaction and expression to accurately prioritize immunogenic neoantigens

**DOI:** 10.1101/2021.06.24.21259393

**Authors:** Elizabeth S. Borden, Kenneth H. Buetow, Bonnie J. LaFleur, Melissa A. Wilson, Karen Taraszka Hastings

## Abstract

Accurate prioritization of immunogenic neoantigens is key to developing personalized cancer vaccines and distinguishing those patients likely to respond to immune checkpoint inhibition. However, there is no consensus regarding which characteristics best predict neoantigen immunogenicity, and no model to date has both high sensitivity and specificity and a significant association with survival in response to immunotherapy. We address these challenges in the prioritization of immunogenic neoantigens by 1) identifying which neoantigen characteristics best predict immunogenicity, 2) integrating these characteristics into an immunogenicity score, NeoScore, and 3) demonstrating an improved association of the NeoScore with response to immune checkpoint inhibition compared to mutational burden. One thousand random and evenly split combinations of immunogenic and non-immunogenic neoantigens from a validated dataset were analyzed using a regularized regression model for characteristic selection. The selected characteristics, the dissociation constant and binding stability of the neoantigen:MHC class I complex and expression of the mutated gene in the tumor, were integrated into the NeoScore. A web application is provided for calculation of the NeoScore. The NeoScore results in improved, or equivalent, performance in four test datasets as measured by sensitivity, specificity, and area under the receiver operator characteristics curve compared to previous models. Among cutaneous melanoma patients treated with immune checkpoint inhibition, a high NeoScore had a greater association with improved survival compared to mutational burden. Overall, the NeoScore has the potential to improve neoantigen prioritization for the development of personalized vaccines and contribute to the determination of which patients are likely to respond to immunotherapy.

## Introduction

Cancers arise through mutations in the genome of healthy human cells. As these mutations occur, some will produce mutated proteins, which have the potential to be processed into neoantigens that bind MHC class I and are presented on the cell surface. These neoantigens then act as tumor-specific targets with the potential to elicit a cytotoxic CD8+ T cell response (Tran, Robbins, and Rosenberg 2017; Schumacher and Schreiber 2015; Yarchoan *et al*. 2017; J. P. Ward, Gubin, and Schreiber 2016). Tumor-specific neoantigens have strong potential to be targets of T cell-mediated destruction, because they are not subject to immune tolerance or non-reactivity to self. However, there are two key ways in which the above mechanism may fail in tumor destruction. For one, recent evidence suggests that most neoantigens do not elicit an immune response in their natural state, termed immunologic ignorance (Linette *et al*. 2019). Second, once a T cell response is mounted to a neoantigen, that response may become exhausted over time due to inhibitory signals from the tumor microenvironment (Rausch and Hastings 2017).

Circumventing these limitations to re-invigorate the host immune response has been the goal of many recent cancer therapies. To address immunologic ignorance, personalized cancer vaccines have been created and have demonstrated early success (Sahin *et al*. 2017; Hilf *et al*. 2019; Keskin *et al*. 2019; Carreno *et al*. 2015; Gubin *et al*. 2015; Ott *et al*. 2017). These vaccines have taken several forms, including direct exposure to neoantigens (Ott *et al*. 2017), neoantigen-encoding RNA vaccines (Sahin *et al*. 2017), neoantigen-loaded dendritic cell vaccines (Carreno *et al*. 2015), and adoptive transfer of neoantigen-specific T cells (Tran *et al*. 2014; Yee *et al*. 2002; Zacharakis *et al*. 2018). Each of these methods requires accurate knowledge of the neoantigens presented by the tumor cell with the potential to elicit an immune response. *In silico* prioritization methods have been used to prioritize which set of neoantigens should be experimentally tested, but the ability to prioritize the immunogenicity of each neoantigen, with high sensitivity and specificity, is still limited (Wells *et al*. 2020; Zhou *et al*. 2019; Wood *et al*. 2018; Bjerregaard *et al*. 2017; Kim *et al*. 2018).

Exhaustion of T cells and attenuation of T cell activation can be overcome by immune checkpoint inhibition, such as monoclonal antibodies against PD-1 or CTLA-4. Immune checkpoint inhibition blocks inhibitory signals to the T cells to enhance T cell-mediated tumor destruction (Gubin *et al*. 2014; Van Allen *et al*. 2015; Lommatzsch, Bratke, and Stoll 2018). However, immune checkpoint inhibition is only effective in a subset of patients (Rausch and Hastings 2017), and there is no consensus on how to prioritize which patients will respond (McGranahan *et al*. 2016; Hellmann *et al*. 2019; Rizvi, Hellmann, *et al*. 2015; Van Allen *et al*. 2015; Snyder *et al*. 2014; Łuksza *et al*. 2017; Zhou *et al*. 2019). In a pan-cancer analysis, Yarchoan *et al*. demonstrated that cancer types with a higher mutational burden, such as melanoma, had improved response to anti-PD-1 therapy compared to cancer types with a lower mutational burden (Yarchoan, Hopkins, and Jaffee 2017). There are limitations to mutational burden as a predictor of response to immune checkpoint inhibition.

First, in multiple myeloma, there was an association of increased tumor mutational burden with decreased response to immune checkpoint inhibition (Miller *et al*. 2017). Second, in melanoma, the association between the tumor mutational burden and response to immune checkpoint inhibition was confounded by the melanoma subtype (Liu *et al*. 2019). Finally, in lung cancers that progressed after treatment with immune checkpoint inhibitors, there was an increase in the tumor mutational burden compared to the pretreatment state (Anagnostou *et al*. 2017), thus, contradicting the expectation that tumor cells resistant to immune checkpoint inhibition would have a low number of neoantigens. Together, these findings suggest that tumor mutational burden is not sufficient for predicting response to immune checkpoint inhibition.

Several recent papers have been dedicated to predicting neoantigen immunogenicity based on the characteristics of validated immunogenic neoantigens (Zhou *et al*. 2019; Łuksza *et al*. 2017; Wood *et al*. 2018; Wells *et al*. 2020; Schmidt *et al*. 2020). However, there is no consensus regarding which neoantigen characteristics are important for the prioritization of immunogenic neoantigens or the best way to combine the characteristics into an overall immunogenicity score. To identify the characteristics that best prioritize the immunogenicity of the neoantigens, a model-based approach was applied to evaluate neoantigen characteristics encompassing expression, processing, presentation, and T cell recognition in prioritizing neoantigen immunogenicity. The selected characteristics were then combined into an overall logistic regression model called the “NeoScore.” The development of the NeoScore has largely focused on melanoma, except for one lung cancer patient included in the TESLA consortium dataset (Wells *et al*. 2020).

Immune checkpoint inhibition and personalized neoantigen vaccines are particularly effective in mutation-rich melanoma (Hodi *et al*. 2018; Rizvi, Garon, *et al*. 2015; Solomon, Beavis, and Darcy 2020; Shemesh *et al*. 2021). However, even in melanoma, a positive outcome from these interventions is not assured (Rausch and Hastings 2017). To assess the clinical utility of the NeoScore and its ability to improve assessment of outcome in melanoma, the relationship of the NeoScore to survival in response to immunotherapy was tested using the datasets of Van Allen *et al*. 2015 and Liu *et al*. 2019.

## Results

### Overlap of Strelka and GATK Mutect2 identifies the maximum number of validated immunogenic mutations

The first step in effective prioritization of neoantigens is to identify a high-fidelity list of somatic mutations. Isolating somatic mutations in cancers is more difficult than variant calling in normal tissue since cancers do not follow the typical rules of copy number or heterozygosity and often consist of multiple clonal populations with normal tissue contamination. The TESLA consortium validated neoantigens derived from mutations identified by 25 teams. The methods used for identifying the somatic mutations were not reported, making it difficult to reproduce all neoantigens (Wells *et al*. 2020). To maximize the immunogenic mutations identified, three highly rated programs were used to identify single nucleotide variants (SNVs) and small insertions and deletions (indels) for the data from the TESLA consortium: VarScan2, GATK Mutect2, and Strelka (Koboldt *et al*. 2012; Benjamin *et al*. 2019; Saunders *et al*. 2012). A large degree of overlap was found in the ability of each program to identify the 34 validated immunogenic mutations. As shown in Figure 1, GATK Mutect2 and Strelka successfully identified 27/34 mutations, while VarScan2 identified 24/34 (Supplementary Tables 1). The mutations from each of these programs overlapped, such that 27 was the maximum number of immunogenic mutations identified. To ensure that the success of VarScan2 in identifying the immunogenic mutations was not hindered by over-filtering, the unfiltered output was assessed and only one immunogenic mutation had been eliminated by filtration steps (Supplementary Table 1). GATK Mutect2 and Strelka identified the same number of immunogenic mutations with or without their respective filters (data not shown). LoFreq was also tested for the identification of somatic mutations, as LoFreq is optimized to call low-frequency mutations (Wilm *et al*. 2012). However, LoFreq did not identify any additional immunogenic mutations (Supplementary Figure 1, Supplementary Table 1). The neoantigens derived from unidentified immunogenic mutations may be due to the reference genome version, peptide generation steps, or mutations identified by programs that were not tested. Based on these results in the TESLA consortium data, the overlap of GATK Mutect2 and Strelka was used in applying our model to predict progression-free survival in response to immune checkpoint inhibition in melanoma. Overall, the combination of GATK Mutect2 and Strelka identified greater than or equal to the number of validated immunogenic neoantigens as in all other reported pipelines (Wells *et al*. 2020), while simultaneously decreasing the total number of potential neoantigens by 89.44%.

**Figure 1.**
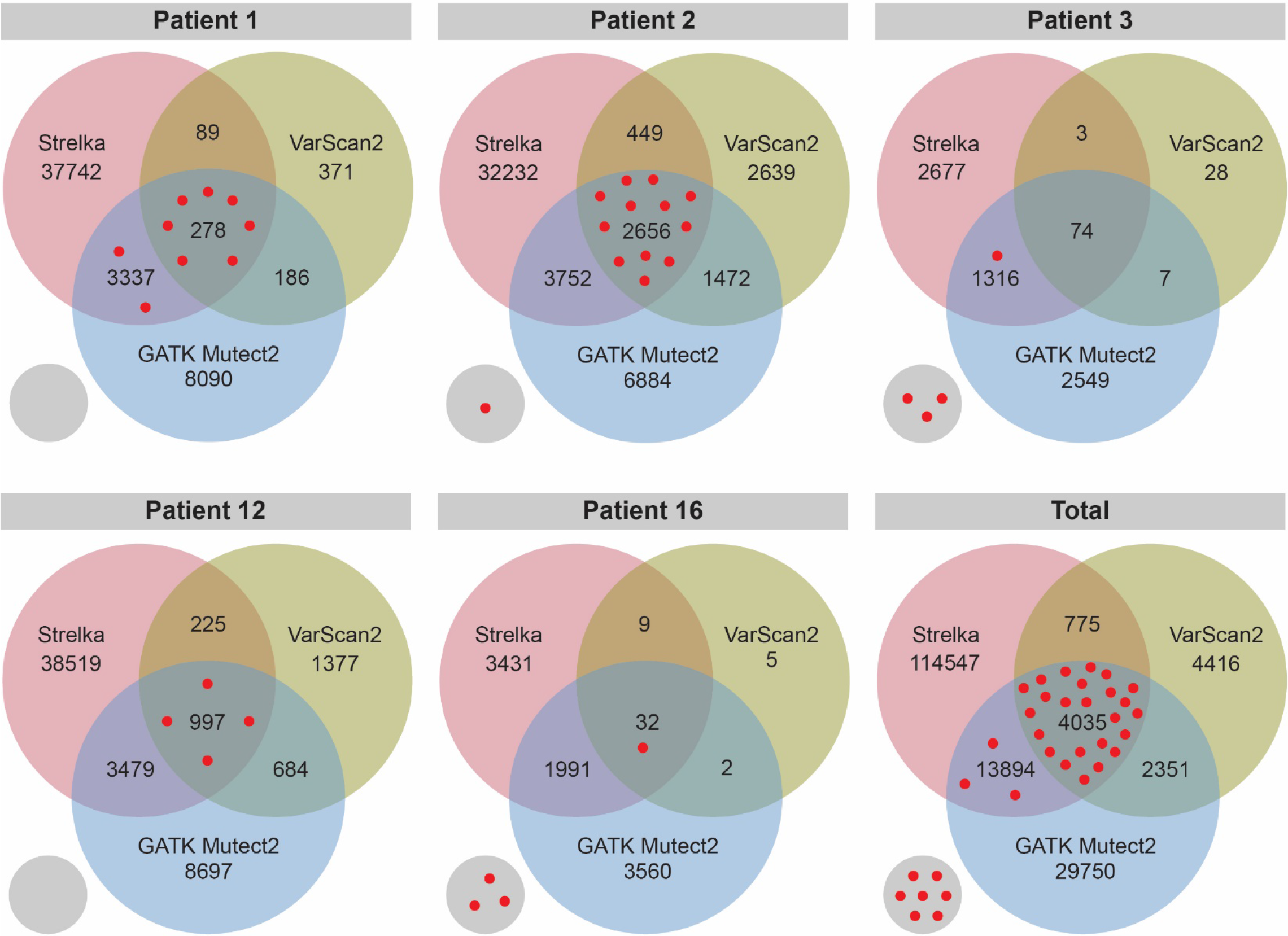
Maximum Number of Validated Immunogenic Mutations Identified by the Overlap of Strelka and GATK Mutect2. Comparison of the number of neoantigens derived from single nucleotide variants (SNVs) and small insertions and deletions (indels) by each of three different programs (Varscan2, GATK Mutect2, and Strelka) in the TESLA consortium dataset. Red dots represent the validated immunogenic neoantigens from each patient. The gray circle represents validated immunogenic neoantigens that were not identified by the three programs.

### Dissociation constant and binding stability of the neoantigen:MHC class I complex as well as expression are significantly different between immunogenic and non-immunogenic neoantigens

A set of computationally predicted neoantigen characteristics were calculated for the expression, processing, presentation, and T cell receptor (TCR) recognition of SNV and small indel-derived neoantigens (Figure 2A). Figure 2B demonstrates that the set of characteristics considered is inclusive of all of those considered by the three models to which the NeoScore is compared. To begin, the distribution of each of these characteristics was assessed for immunogenic and non-immunogenic neoantigens. A log transformation was applied to standardize characteristics with a large degree of skewness. The neoantigens assessed here and used to fit the overall model were restricted by three inclusion criteria: 1) they had been tested for immunogenicity by the TESLA consortium, 2) they were identified by GATK Mutect2 and Strelka, and 3) they had expression data. The final dataset used includes 5 patients with a total of 344 neoantigens, which had been tested for their ability to elicit a T cell response using multimer staining. The TESLA consortium found that 26 of the 344 tested neoantigens elicited a T cell response in an unvaccinated state (Wells *et al*. 2020).

**Figure 2.**
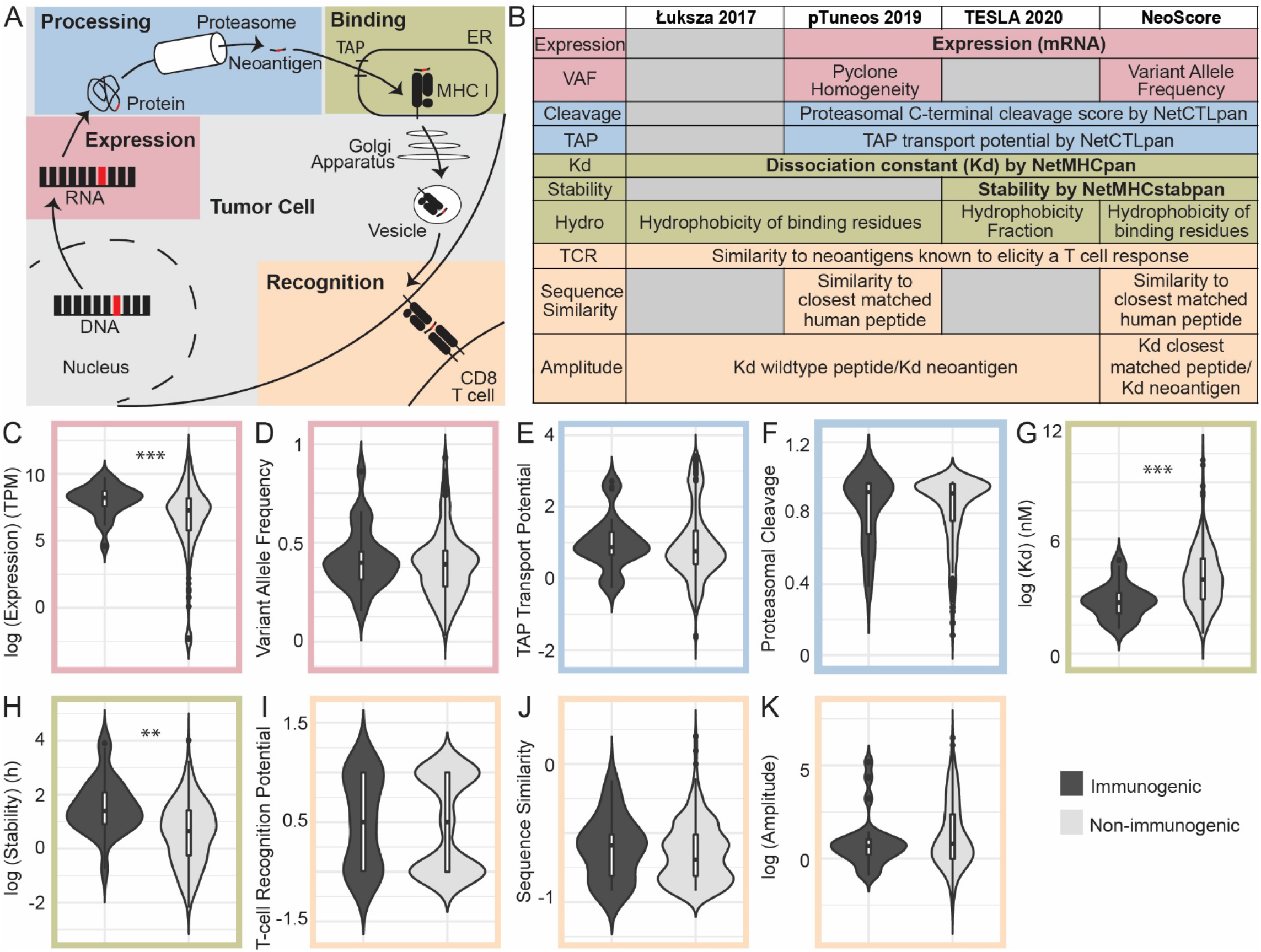
Expression, Dissociation Constant, and Stability are Significantly Different Between Immunogenic and Non-Immunogenic Neoantigens. (A) Steps in the formation of an MHC class I-restricted immunogenic neoantigen. (B) Computationally predicted neoantigen characteristics used to assess the potential for immunogenicity in the NeoScore model in comparison to three other models. Expression consists of the gene-level mRNA expression and clonality or variant allele frequency of the mutation. Processing consists of proteasomal cleavage and TAP transport potential. MHC class I binding is assessed through the MHC class I dissociation constant, binding stability, and hydrophobicity (Figure 3) of the neoantigen. The potential to stimulate a T cell response is assessed by the similarity to known T cell epitopes, the sequence similarity to normal human peptides, and the relative MHC class I dissociation constant compared to the dissociation constant of the closest matched normal peptide. All ten characteristics listed under the NeoScore model were considered for inclusion, and those in bold (expression, dissociation constant, and binding stability) were the selected characteristics for the final model. (C-K) Comparison of the distribution of the characteristics for immunogenic and non-immunogenic neoantigens. (C) Log-transformed distribution of mRNA expression. (D) Distribution of variant allele frequency (VAF) calculated by GATK Mutect2. (E-F) Distribution of proteasomal cleavage and TAP transport potential calculated by NetCTLpan. (G) Log-transformed distribution of the MHC class I to neoantigen dissociation constant calculated by NetMHCpan. (H) Log-transformed distribution of the MHC class I to neoantigen binding stability calculated by NetMHCstabpan. (I) T cell recognition potential calculated using the model from Łuksza et al. (J) Log-transformed distribution of the BLOSUM62 sequence similarity between the closest matched human peptide and the neoantigen of interest and divided by the length of the neoantigen to normalize. (K) Log-transformed distribution of the amplitude calculated as the ratio of the dissociation constant of the closest matched human peptide to MHC class I to the dissociation constant of the neoantigen to MHC class I. **:p<0.001, ***:p<10^−5^

Expression was considered in two ways: 1) mRNA expression level as calculated from RNAseq data and 2) variant allele frequency (VAF). Immunogenic neoantigens had a significantly higher expression level than non-immunogenic neoantigens (p=6.835×10^−6^, Figure 2C). The clonality of the neoantigen was calculated by FastClone (Xiao *et al*. 2020), but FastClone did not converge for 4/6 of the samples, so the VAF was used instead. No significant difference in the VAF was observed between immunogenic and non-immunogenic neoantigens (p=0.360, Figure 2D).

Processing steps for the neoantigen were considered as both the proteasomal cleavage potential and TAP (transporter associated with antigen processing) potential scores from netCTLpan (Stranzl *et al*. 2010). While there was a higher average for both characteristics in the immunogenic than non-immunogenic neoantigens, no statistically significant difference was observed (p=0.858 and p=0.848, respectively) (Figure 2E-F). The binding to the MHC class I molecule was then considered by both the dissociation constant and stability of the MHC class I:neoantigen interaction. The dissociation constant was calculated by netMHCpan (Reynisson *et al*. 2020). The MHC class I dissociation constants were significantly lower in immunogenic than non-immunogenic neoantigens, indicating a higher binding affinity of immunogenic neoantigens to MHC class I (p=2.69×10^−7^, Figure 2G). The binding stability was calculated by netMHCstabpan (Rasmussen *et al*. 2016) and showed significantly higher values for immunogenic over non-immunogenic neoantigens. (p=4.199×10^−5^, Figure 2H).

The ability of the neoantigen to stimulate a T cell response was predicted using the model created by Łuksza *et al*. to calculate a characteristic referred to as the TCR recognition potential (Łuksza *et al*. 2017). The TCR recognition potential is a probabilistic model that considers the sequence similarity between the neoantigen and a known T cell epitope from the Immune Epitope Database (IEDB) as a proxy for the binding affinity of the neoantigen:MHC class I-TCR interaction. The TCR recognition potential showed no statistically significant difference between the immunogenic and non-immunogenic neoantigens (p=0.670, Figure 2I). Two methods were then applied to account for T cell development. During maturation in the thymus, T cells expressing TCRs with high avidity to normal human peptides undergo apoptosis. Thus, a neoantigen with a high degree of sequence similarity to a normal human peptide is less likely to elicit a T cell response. The first method used to account for T cell development was the sequence similarity to the closest matched normal human peptide. The sequence similarity was calculated by performing a BLAST search and subsequently a BLOSUM62 alignment matrix as described (Wood *et al*. 2018). The resulting sequence similarity was normalized by dividing by the neoantigen length. No statistically significant difference was observed in the sequence similarity for immunogenic and non-immunogenic neoantigens (p=0.206, Figure 2J). The second method considered was the amplitude. The amplitude is calculated by adjusting the dissociation constant of the neoantigen:MHC class I complex by the dissociation constant of the closest matched human peptide:MHC class I complex. The amplitude adjusts for the regulation of TCR specificities during T cell maturation but also considers that only a normal human peptide capable of binding an MHC class I molecule will significantly impact the immunogenicity of the neoantigen. The amplitude was not significantly different between immunogenic and non-immunogenic neoantigens (p=0.206, Figure 2K).

One final characteristic considered was the hydrophobicity of the neoantigen. The hydrophobicity of the neoantigen has been proposed to be associated with greater neoantigen immunogenicity because of the hydrophobicity of key anchor residues in the MHC class I binding cleft, as well as residues in the TCR, having the greatest interaction with the neoantigen (Chowell *et al*. 2015). Mixed results have been reported for the association of hydrophobicity and immunogenicity to date (Chowell *et al*. 2015; Zhou *et al*. 2019; Łuksza *et al*. 2017; Wells *et al*. 2020). One reason is the use of different methods for hydrophobicity, three of which are considered here. In the first method, the hydrophobicity is calculated as a fraction of the neoantigen residues that are hydrophobic (Wells *et al*. 2020). The TESLA consortium reported a significantly lower hydrophobicity of immunogenic neoantigens compared to non-immunogenic neoantigens, which is in the opposite direction as expected. While there was not a statistically significant difference in the neoantigens included in our analysis, the hydrophobicity fraction still had a lower average for immunogenic than non-immunogenic neoantigens (Figure 3, p=0.211). No difference in hydrophobicity was seen in additional datasets from Carreno or Strønen *et al*., and a significantly higher hydrophobicity of immunogenic neoantigens was observed in the dataset from Ott *et al*. (Figure 3, p=0.0168). The second method calculated the hydrophobicity fraction using the empirical observations from Chowell *et al*. to determine which amino acids would increase the likelihood of immunogenicity (Chowell *et al*. 2015). The method using the observations from Chowell *et al*. considers both hydrophobicity and other chemical properties such as side chain bulkiness and polarity. No differences in hydrophobicity were seen across the four datasets using the empirical observations from Chowell *et al*. (Figure 3). The final method considered the hydrophobicity at the anchor residues. A mutation that changed a previously hydrophobic anchor residue to a hydrophilic residue was given a score of zero, while all other changes or no change were given a score of one (Łuksza *et al*. 2017). While there was no statistically significant difference in hydrophobicity for any of the four datasets using the method of Łuksza et al., three out of four datasets showed a greater percentage of immunogenic neoantigens without a loss of hydrophobicity at an anchor residue (Figure 3). Only the method from Łuksza was included moving forward, since it had the greatest consistency across datasets.

**Figure 3.**
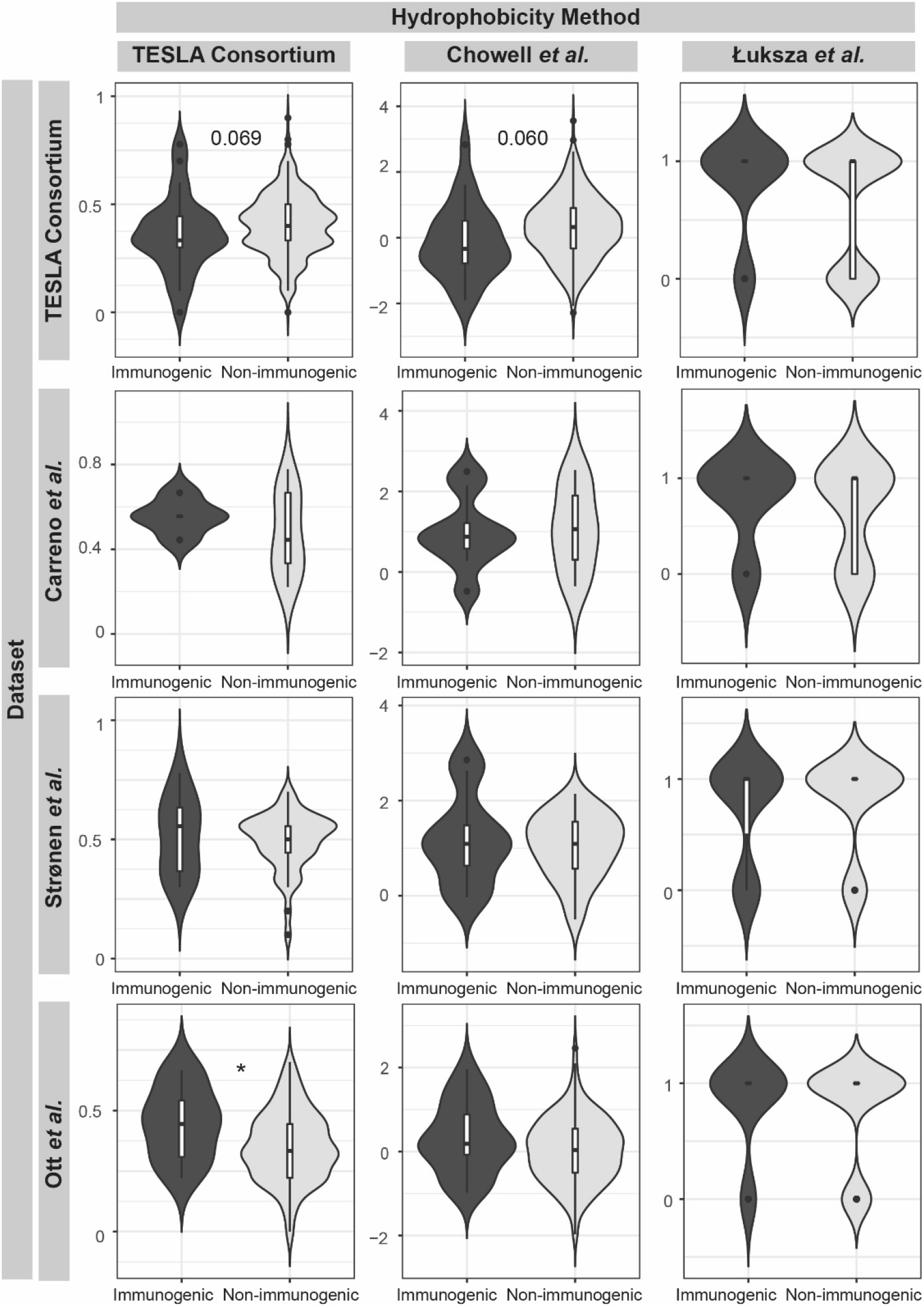
No Method for Calculating Neoantigen Hydrophobicity is Consistently Associated with Immunogenicity. Comparison of the distribution of hydrophobicity values for the TESLA consortium, Carreno, Strønen, and Ott datasets between immunogenic and non-immunogenic neoantigens. Hydrophobicity was calculated by three methods. The TESLA consortium method is calculated as the number of hydrophobic amino acids divided by the total number of amino acids. The method based on the Chowell et al. empirical data uses the same calculation scheme but considers both hydrophobicity and other chemical properties such as side chain bulkiness and polarity in ranking the neoantigens. The Łuksza et al. method considers the change of an amino acid at the anchor residues compared to the closest matched human peptide. *:p<0.05

Next, the degree of correlation between the characteristics calculated for each neoantigen was assessed. Only two characteristics were significantly correlated: the dissociation constant and the binding stability (Figure 4A). Despite their correlation, there is evidence that the two characteristics both contribute to accurate neoantigen prioritization. While the dissociation constant assesses the affinity of the interaction between the neoantigen and the MHC class I molecule, the stability predicts the length of time that the neoantigen will remain bound. The importance of binding stability is demonstrated in Figure 4B, where clustering of the immunogenic neoantigens in the upper left-hand corner can be observed, indicating the influence of both characteristics in determining immunogenicity.

**Figure 4.**
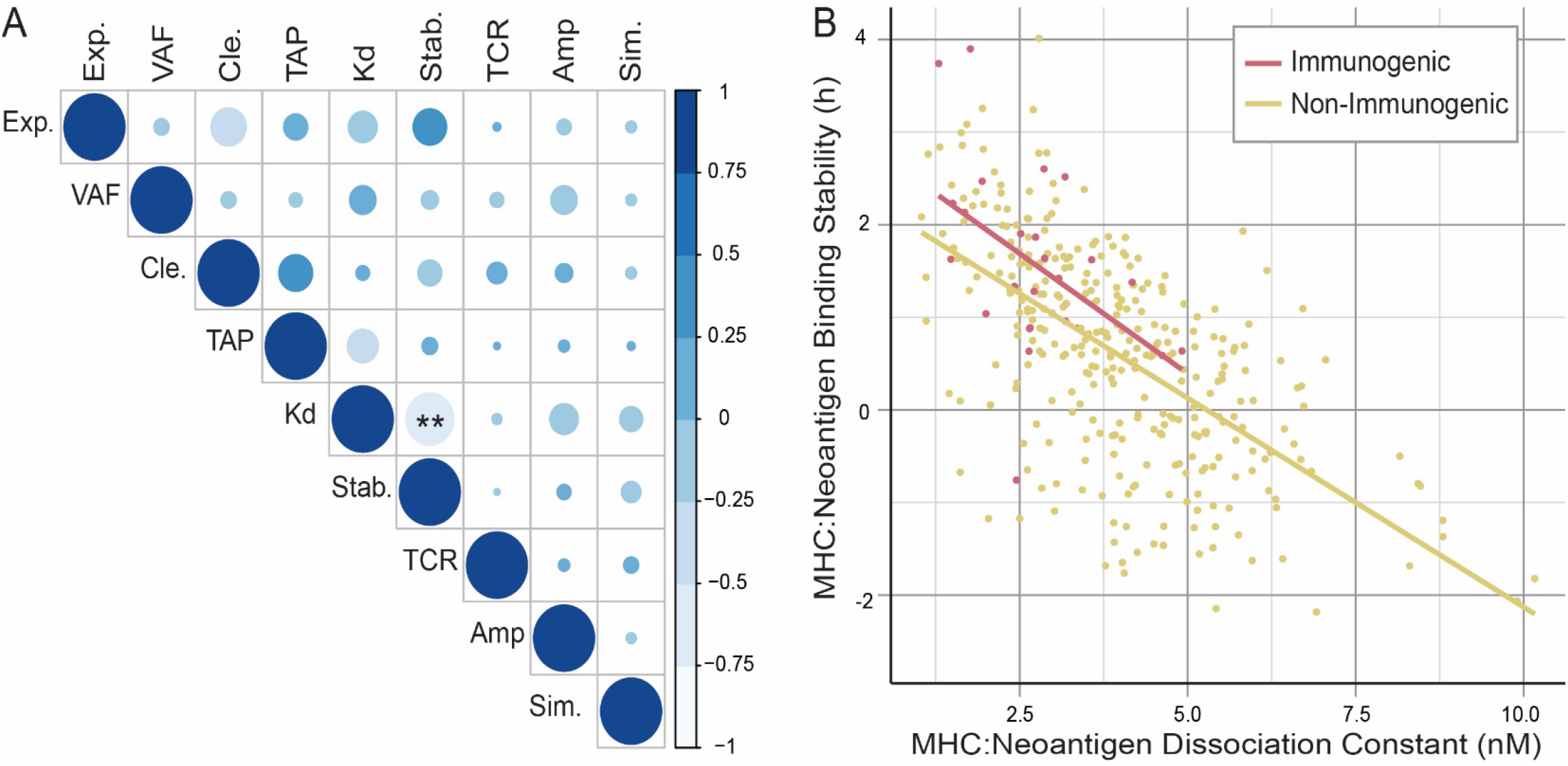
Binding Stability and Dissociation Constant are Significantly Correlated but Both Contribute to Immunogenicity. (A) Correlation between all calculated neoantigen characteristics: mRNA expression (Exp.), Variant Allele Frequency (VAF), proteasomal cleavage potential (Cle.), TAP transport potential (TAP), MHC class I to neoantigen dissociation constant (Kd), MHC class I to neoantigen binding stability (Stab.), T cell recognition potential (TCR), amplitude, defined as the ratio of the MHC class I binding of the closest matched human peptide and the neoantigen (Amp.), and normalized sequence similarity score for closest matched human peptide and the neoantigen (Sim.). (B) Correlation of MHC class I dissociation constant with MHC class I binding stability of the neoantigen. Lines show the linear relationship between the characteristics for immunogenic (red) and non-immunogenic (yellow) neoantigens, respectively. **:p<0.001

Capietto *et al*. recently found that a different set of neoantigen characteristics will influence immunogenicity, if the mutation occurs in an anchor residue compared to mutations in non-anchor residues (Capietto *et al*. 2020). They demonstrated that, if a mutation occurred in an anchor residue, the amplitude had a greater predictive value than the dissociation constant of the neoantigen:MHC class I complex alone. To assess the influence of the mutation position, the distribution of the amplitude and the unadjusted dissociation constant for neoantigens with a mutation in an anchor or a non-anchor residue were analyzed. To maximize the chances of detecting a significant difference with the relatively low number of immunogenic neoantigens derived from mutations in anchor residues (n=13), the analysis of the impact of mutation’s position was performed across the combination of four datasets (Strønen *et al*. 2016; Ott *et al*. 2017; Carreno *et al*. 2015; Wells *et al*. 2020). As shown in Supplementary Figure 2, no statistically significant difference was observed for the amplitude with either anchor or non-anchor residue mutations. While the anchor-residue mutations did have a higher average amplitude in immunogenic neoantigens, the difference was not statistically significant. In contrast, the dissociation constant was significantly lower in immunogenic neoantigens than non-immunogenic neoantigens. Therefore, there was not a compelling reason to fit separate models for neoantigens with mutations in anchor residues and those with mutations in non-anchor residues. Furthermore, because the amplitude is mathematically dependent on the dissociation constant, the amplitude was not included moving forward.

### Regularized regression approach creates a neoantigen prioritization model, NeoScore, that outperforms existing models that score each neoantigen

While analysis of the distribution of each characteristic determined those with a statistically significant difference between immunogenic and non-immunogenic neoantigens, we next assessed whether other characteristics influenced the ability to optimally prioritize SNV and small indel-derived immunogenic neoantigens. A regularized regression is an independent, model-based approach to determine the group of characteristics that are important for discriminating between immunogenic and non-immunogenic neoantigens, whether or not each characteristic is statistically significant. A logistic model using an elastic net-based regularization was unable to identify individual characteristics that were predictive of immunogenicity after shrinkage penalties were applied; a consequence that is often seen with a small effective sample size. The effective sample size for a binomial analysis is based on the class with the smallest number of observations, which is the immunogenic neoantigens (n=26). Consequently, a cross-validation approach was applied to select the best subset of neoantigen characteristics. One thousand combinations of 26 non-immunogenic and 26 immunogenic neoantigens were randomly selected, and the elastic net regularized regression was fit on each combination (Figure 5A). Performing 1000 random samples allowed for the examination of the impact of neoantigen characteristics on immunogenicity while adjusting for the small effective sample size. The number of times each characteristic was selected out of the 1000 random combinations was tracked. The dissociation constant, binding stability, and mRNA expression were each selected over 800 of the 1000 times, while no other characteristic was selected over 500 times (Figure 5B). These fits were consistently able to distinguish immunogenic and non-immunogenic neoantigens, as demonstrated by the high density of area under the receiver operator characteristics curve (AUC) values around the mean of 0.85 (25th percentile 0.818, 75th percentile 0.885) (Figure 5C).

**Figure 5.**
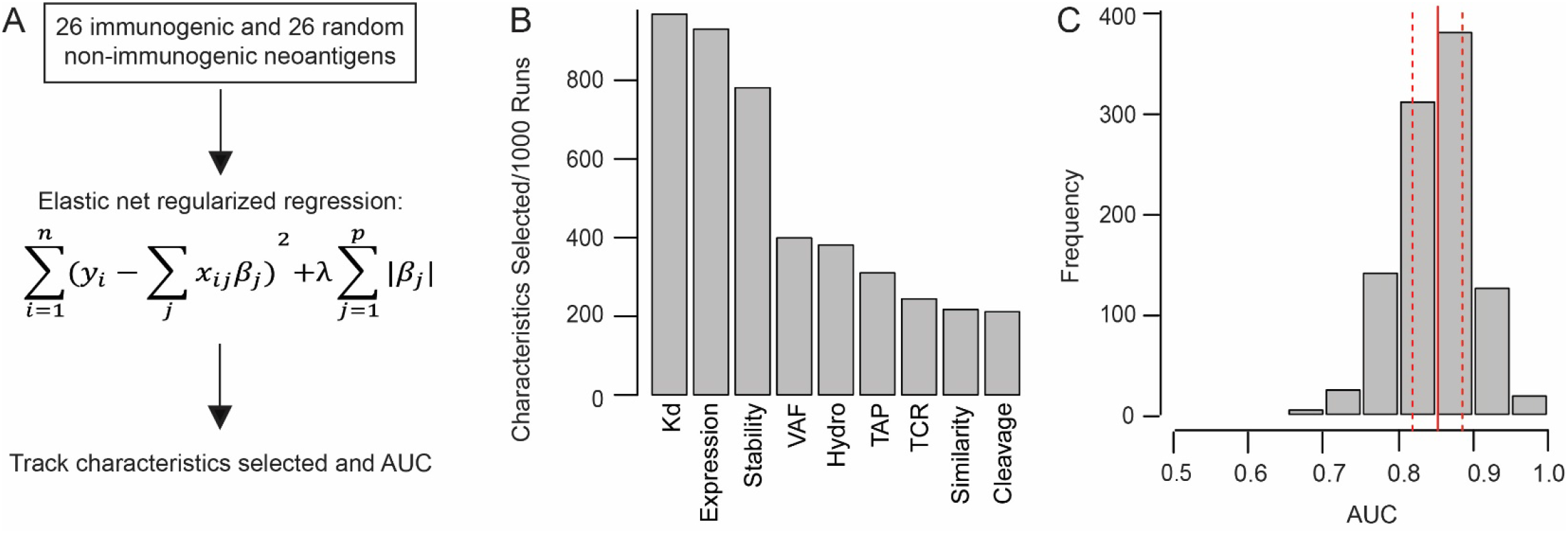
Regularized Regression Approach Selects Dissociation Constant, Expression, and Stability as the Characteristics of Greatest Importance in Prioritizing Immunogenicity. Regularized (elastic net) regression approach to the selection of neoantigen characteristics. (A) Process used for characteristic selection. 26 immunogenic and 26 randomly selected non-immunogenic neoantigens were isolated 1000 times and fit with an elastic net regression. For every fit, the characteristics selected by the model as well as its performance were tracked. (B) Number of times each characteristic was selected out of 1000 runs. The characteristics included are as follows: MHC class I to neoantigen dissociation constant (Kd), mRNA expression (Expression), MHC class I to neoantigen binding stability (Stability), Variant Allele Frequency (VAF), hydrophobicity of the anchor residues (Hydro), TAP transport potential (TAP), T cell recognition potential (TCR), normalized sequence similarity score for closest matched human peptide and the neoantigen (Similarity), and proteasomal cleavage potential (Cleavage). (C) Distribution of the area under the receiver operator characteristics curve (AUC) values for each of the 1000 fits. The solid red line indicates the mean AUC, and the dashed red lines represent the 25th and 75th percentile, respectively.

Based on the results of the model-based regression approach, a logistic regression model was fit with the dissociation constant, binding stability, and expression in the TESLA consortium data. The final logistic regression model will be called the “NeoScore.” The equation for the model is as follows:

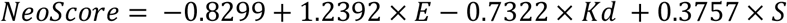

Where *E* is the scaled, log-transformed expression value, *Kd* is the log-transformed dissociation constant in units of nanomolar (nM), and *S* is the log-transformed stability measured as the half-life of the binding interaction in units of hours. The coefficients for expression and stability were both positive, as expected since higher expression and longer binding times likely increase the chance of a neoantigen to elicit an immune response. The coefficient for the dissociation constant was negative, as expected since a lower dissociation constant indicates a higher binding affinity. These coefficients match the observed directions of change from Figure 2. Raw data from NetMHCpan, NetMHCstabpan, and Salmon can be processed and combined to return a set of neoantigens prioritized by their NeoScore using the following web application: https://bordene.shinyapps.io/MHCI_neoantigen_prioritization/.

In the TESLA consortium dataset, the NeoScore had an AUC of 0.846, which exceeds the AUC of 0.70 needed to be considered a discriminatory model (Hosmer, Lemeshow, and Sturdivant 2013). The NeoScore also outperformed the AUC of the Łuksza model (AUC=0.615) (Table 1; Figure 6A). The NeoScore was then tested in four additional datasets. The NeoScore outperformed the Łuksza model in the Cohen (0.834 AUC vs. 0.689 AUC) and Strønen datasets (0.739 AUC vs. 0.620 AUC) (Cohen *et al*. 2015; Strønen *et al*. 2016) (Table 1; Figure 6B,C). In the Carreno dataset, the NeoScore slightly outperformed Łuksza and the pTuneos hydrophobicity model (0.704 AUC for NeoScore, 0.696 AUC for pTuneos hydrophobicity, and 0.657 AUC for Łuksza) (Table 1; Figure 6D). Published results of the immunogenicity scores from the pTuneos model were used (Zhou *et al*. 2019), as the model was not able to be successfully run with the other datasets. Both the hydrophobicity-only model and the full model provided by pTuneos are included, as the hydrophobicity model outperformed the full model. Similarly, in the Ott dataset, the NeoScore slightly outperformed the Łuksza model (0.609 AUC for NeoScore and 0.575 AUC for Łuksza) (Ott *et al*. 2017) (Table 1; Figure 6E).

**Table 1:** Comparison of the AUC and optimal threshold sensitivities and specificities for the prediction of immunogenic neoantigens using NeoScore vs. other models. The abbreviated NeoScore omits expression.

| Dataset | Model | Sensitivity [95% C.I.] | Specificity [95% C.I.] | AUC |
| --- | --- | --- | --- | --- |
| TESLA consortium (n = 344) | Łuksza | 0.692 [0.385-0.885] | 0.561 [0.433-0.815] | 0.615 |
|  | TESLA consortium | 0.808 | 0.692 | ----- |
|  | **NeoScore** | **0.846 [0.730-1.00]** | **0.758 [0.550-0.887]** | **0.846** |
|  | **Abbreviated NeoScore** | **0.846 [0.731-1.00]** | **0.638 [0.522-0.755]** | **0.770** |
| Cohen (n = 357) | Łuksza | 1.00 [0.571-1.00] | 0.394 [0.354-0.871] | 0.689 |
|  | TESLA consortium | 0.857 | 0.631 | ----- |
|  | **NeoScore** | **0.857 [0.571-1.00]** | **0.566 [0.514-0.614]** | **0.834** |
|  | **Abbreviated NeoScore** | **0.722 [0.500-0.944]** | **0.347 [0.265-0.429]** | **0.722** |
| Strønen (n = 57) | Łuksza | 0.727 [0.455-1.00] | 0.422 [0.289-0.556] | 0.620 |
|  | TESLA consortium | 0.727 | 0.733 | ----- |
|  | **NeoScore** | **0.455 [0.182-0.727]** | **0.822 [0.711-0.933]** | **0.739** |
|  | **Abbreviated NeoScore** | **0.818 [0.545-1.00]** | **0.778 [0.667-0.889]** | **0.792** |
| Carreno (n = 21) | Łuksza | 0.778 [0.553-1.00] | 0.583 [0.333-0.833] | 0.657 |
|  | TESLA consortium | 0.556 | 0.750 | ----- |
|  | pTuneos hydrophobicity | 0.5 [0.125-1.00] | 0.857 [0.429-1.00] | 0.696 |
|  | pTuneos full model | 0.250 [0.122-1.00] | 1.00 [0.143-1.00] | 0.546 |
|  | **NeoScore** | **0.444 [0.111-0.778]** | **0.833 [0.583-1.00]** | **0.704** |
|  | **Abbreviated NeoScore** | **1.00 [1.00-1.00]** | **0.750 [0.5-1.00]** | **0.935** |
| **Ott (n = 165)** | Łuksza | 0.680 [0.088-0.755] | 0.500 [0.333-1.00] | 0.575 |
|  | TESLA consortium | 0.389 | 0.850 | ----- |
|  | **NeoScore** | **0.278 [0.056-0.500]** | **0.857 [0.796-0.912]** | **0.609** |
|  | **Abbreviated NeoScore** | **0.389 [0.167-0.611]** | **0.830 [0.762-0.885]** | **0.596** |

**Figure 6.**
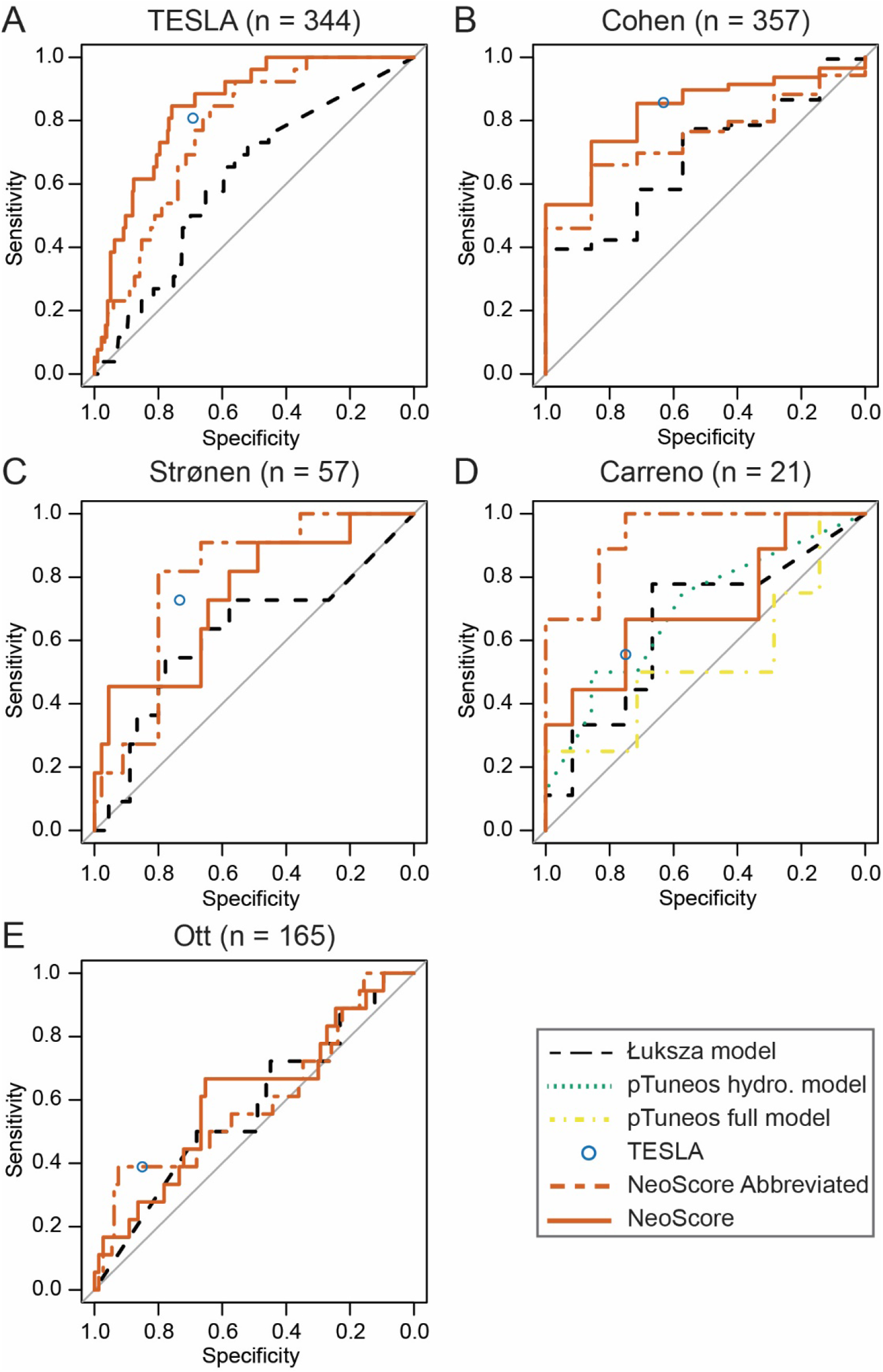
NeoScore Outperforms the Model by Łuksza and Performs Equivalently to the Model by the TESLA Consortium. Comparison of the area under the receiver operator characteristics curves (AUC) for the NeoScore and abbreviated NeoScore compared to the models by Łuksza, the TESLA consortium, and pTuneos. A) Performance of the NeoScore and abbreviated NeoScore in the TESLA consortium dataset (the dataset in which the model was fit). Comparison of the NeoScore and abbreviated NeoScore with existing models in the (B) Cohen dataset, (C) Strønen dataset, (D) Carreno dataset, and (E) Ott dataset.

Since the model by the TESLA consortium consists of a single set of recommended thresholds for the neoantigen:MHC class I dissociation constant, neoantigen:MHC class I binding stability, and expression, the NeoScore could not be compared to the TESLA consortium model in terms of the AUC. Therefore, an optimal threshold for the NeoScore was selected using optimal cutpointr from R statistical software (https://github.com/thie1e/cutpointr) that maximized the sum of the sensitivity and specificity in the TESLA dataset and classified the NeoScore into high and low immunogenicity. The reported sensitivity and specificity for each dataset are based on the threshold optimized in the TESLA dataset (−2.3346). Across all datasets considered, the sensitivity and specificity of the NeoScore were statistically equivalent to that achieved by the TESLA consortium, as demonstrated by the overlap of the 95% confidence interval (Table 1).

Despite the high performance in the TESLA and Cohen datasets, there is a marked decrease in the performance of the NeoScore in the Carreno, Strønen, and Ott datasets. A likely cause for the decreased performance is how the immunogenicity was tested in these datasets. TESLA and Cohen both tested for reactive T cells present in the patient with no additional T cell stimulation. In contrast, Carreno *et al*. administered a dendritic cell vaccine with each of the predicted neoantigens and subsequently tested for an immune response to each neoantigen (Carreno *et al*. 2015). Strønen *et al*. exposed PBMCs from healthy patients to dendritic cells transfected with the neoantigen of interest. They then tested for an immune response to those neoantigens (Strønen *et al*. 2016). Ott *et al*. immunized patients with pools of long synthetic peptides and then tested for an immune response to each neoantigen that could be generated from the long peptides (Ott *et al*. 2017). None of these three methods rely on the expression of the neoantigen in the tumor to activate neoantigen-specific T cells. Therefore, a logistic regression model was fit to the TESLA dataset using only the neoantigen:MHC class I binding stability and dissociation constant. The following abbreviated NeoScore model was obtained:

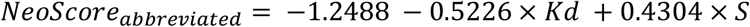

The coefficients obtained for stability and dissociation constant are comparable to those obtained in the full NeoScore model. The threshold for the abbreviated NeoScore was optimized in the TESLA dataset (−2.6269) and then tested in the Cohen, Strønen, Carreno, and Ott datasets. As expected, the abbreviated NeoScore underperformed compared to the full NeoScore model in Cohen and TESLA, but outperformed the full NeoScore model in both Carreno and Strønen (Table 1; Figure 6). These results suggest that expression predicts neoantigen immunogenicity when priming of T cell responses is dependent on expression of the neoantigen by the tumor. However, when the T cells are stimulated independently of expression by the tumor, the predictive benefit of expression is undermined. The poor performance of Ott across models remains unexplained. The Ott dataset did not significantly differ from the other datasets in terms of the distribution of the location of the mutations (anchor vs. non-anchor residues) or the general distribution of the characteristics (data not shown). Overall, elimination of expression enhanced the performance of the model when considering the immune response stimulated independently of the tumor.

To further reaffirm the subset of neoantigen characteristics selected, a logistic regression model was fit using all nine neoantigen characteristics from Figure 5B, which did not notably improve the AUC (Supplementary Figure 3). Furthermore, the optimism, as calculated by the RMS package in R, is much higher for the model that included all neoantigen characteristics (8.20%) than the NeoScore (1.78%). The optimism indicates the likelihood that the model is overfitting the training data, which would make a less generalizable model. The lack of benefit from the additional characteristics adds support that the subset of characteristics selected is optimal for prioritizing immunogenic neoantigens. Overall, the model with the neoantigen:MHC class I dissociation constant and binding stability, and mRNA expression showed the best potential to consistently separate immunogenic and non-immunogenic neoantigens in validated datasets.

### A high NeoScore has a greater association with improved survival compared with mutational burden in cutaneous melanoma patients treated with immunotherapy

Once the ability of the NeoScore to discriminate immunogenic from non-immunogenic neoantigens with high sensitivity and specificity was established, the model was applied to assess clinical outcome in melanoma as measured by progression-free survival in response to immune checkpoint inhibition. The survival analysis was performed across two datasets, one with treatment with anti-CTLA-4 monoclonal antibodies (Van Allen *et al*. 2015) and the other with anti-PD-1 monoclonal antibodies (Liu *et al*. 2019). In both datasets, the cohort was restricted to immunotherapy-naive patients with cutaneous melanoma (Liu *et al*. 2019; Van Allen *et al*. 2015), which allowed us to assess the factors that drive response to immune checkpoint inhibition in melanoma with a high tumor mutational burden and no previous immunoediting. While the range of mutational burden in cutaneous melanoma is wide (1,368-33,591 for the Van Allen dataset and 864-24,292 for the Liu dataset), all samples have a high mutational burden due to UV-induced, DNA mutations. The final cohort sizes were 34 for Van Allen and 53 for Liu. However, the statistical power of the survival analysis is limited by the number of deaths observed in each dataset, resulting in an effective sample size of 22 for Van Allen and 20 for Liu.

Identification of somatic mutations, generation of neoantigens, as well as calculation of expression, neoantigen:MHC class I dissociation constant and binding stability for each of the patients was performed as described in the methods. These three characteristics were then used to calculate the NeoScore for each neoantigen as described above. Since each patient has many neoantigens with a wide range of NeoScores, there are many potential ways to summarize the neoantigen profile for the sake of comparison between patients. Three ways were selected here: 1) the mutational burden, 2) the neoantigen burden, and 3) the highest NeoScore for a neoantigen from the patient, referred to as the “maximum NeoScore.” The mutational burden and neoantigen burden were attempted for the sake of comparison to the literature, while the maximum NeoScore was used in accordance with the principle that a small number of the most immunogenic neoantigens can drive the immune response. Survival analysis was then performed separately for the mutational burden, neoantigen burden, and the maximum NeoScore.

Although an optimal NeoScore threshold was determined for the prediction of neoantigen immunogenicity, when applied as the threshold for the survival analysis there was not a significant association between a maximum NeoScore that exceeded the optimized threshold and increased progression-free survival (data not shown). There are two possible reasons that there is not a significant association at the threshold optimized in the TESLA dataset. First, the optimal threshold was developed using a validated dataset that only included a subset of possible neoantigens, but here the clinical applicability is assessed on a full set of neoantigens. Second, the NeoScore was trained on the natural T cell responses in the absence of any treatment, but patients in the survival datasets underwent treatment with different immunotherapies. Treatment with immunotherapy may increase the immune response to the tumor and decrease the threshold for immunogenicity required to elicit a response, so unique optimal thresholds for survival analysis were determined for the maximum NeoScore using the MaxStat package from R statistical software (Hothorn 2007). MaxStat assesses both the distribution of the variable (here the maximum NeoScore) and the survival across patients to select a threshold that maximizes the difference in the survival between the groups with a high and low NeoScore. Since MaxStat optimizes the threshold to detect a difference in survival, the results of the survival analysis with the optimized thresholds will need to be confirmed in an independent test dataset.

Next, the association of mutational burden and neoantigen burden with survival in response to immune checkpoint inhibition was evaluated. For the sake of consistency, MaxStat was also used to select the optimal threshold for the mutational burden and neoantigen burden. In the Liu dataset, there was no association between mutational burden and progression-free survival in response to immune checkpoint inhibition (p=0.37) (Figure 7A). In the Van Allen dataset, a high tumor mutational burden was significantly associated with poor progression-free survival (p=0.047) (Figure 7B), which is consistent with the findings of Liu *et al*. 2019. The neoantigen burden was strongly correlated with the mutational burden in both datasets (Figure 7C and D). On survival analysis, the same results were observed for neoantigen burden as for mutational burden where a high neoantigen burden was not associated with improved progression-free survival in the Van Allen dataset and was associated with decreased progression-free survival in the Liu dataset (data not shown).

**Figure 7.**
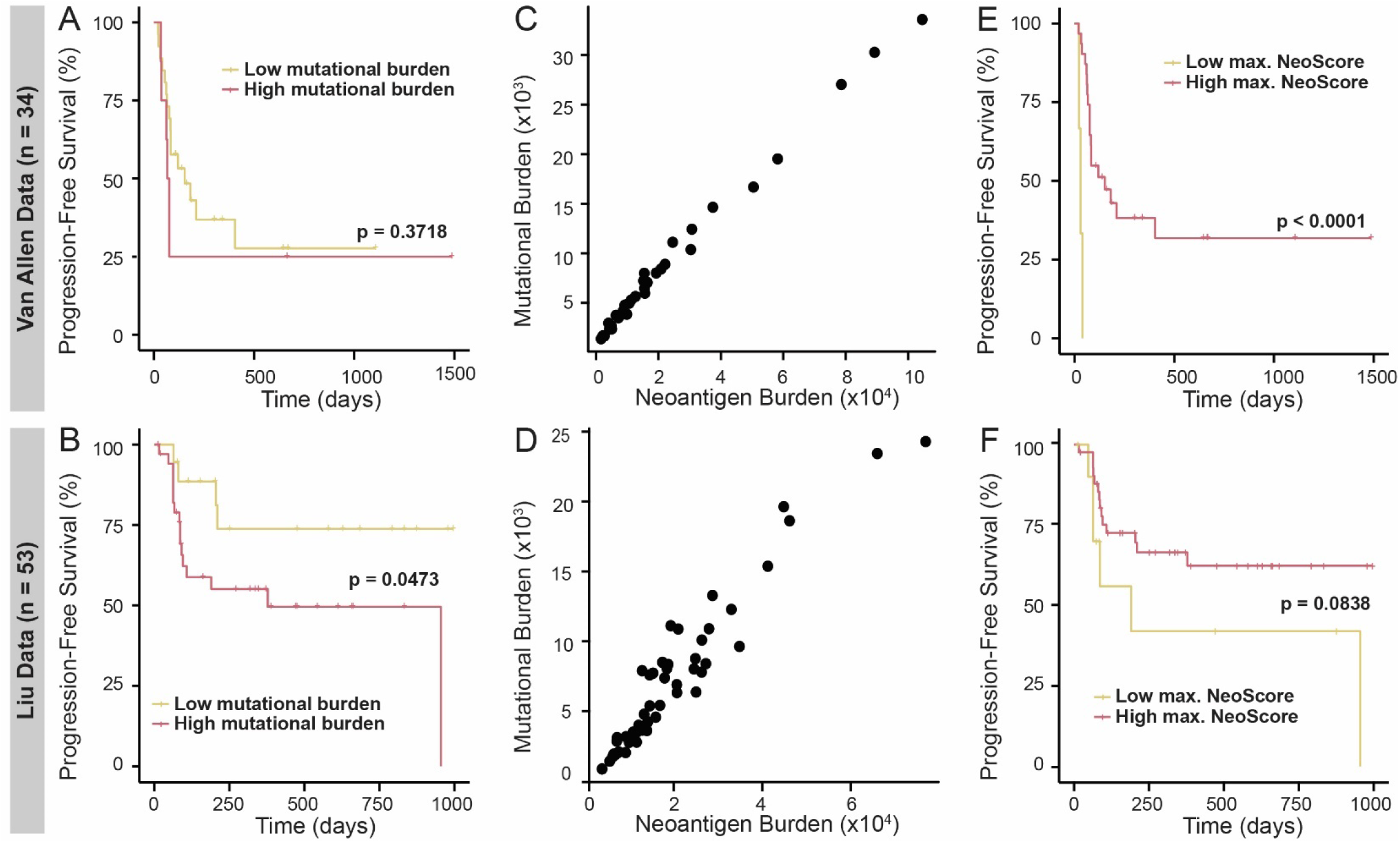
High Maximum NeoScore has Improved Association with Survival Compared to Mutational Burden. Comparison of progression-free survival probability within treatment-naive, cutaneous melanoma patients. All splits between the groups were determined using MaxStat and p-values were calculated using a log-rank test. A high tumor mutational burden is (A) not significantly associated with survival in the Van Allen data and (B) significantly associated with decreased survival in the Liu data. Mutational burden directly correlates with neoantigen burden in (C) the Van Allen data and (D) the Liu data. Patients with a high maximum (max.) NeoScore have (E) significantly increased survival in the Van Allen data and (F) higher survival in the Liu data, but the difference is not statistically significant.

When tested with the MaxStat split, patients in the Van Allen dataset with a high maximum NeoScore (above -2.77) had significantly improved progression-free survival (p < 0.0001) (Figure 7E). Similarly, in the Liu dataset, patients with a high maximum NeoScore (above -2.47) had longer progression-free survival, but the difference was not statistically significant (p=0.084) (Figure 7F). Despite the lack of a statistically significant difference, both datasets show that progression-free survival is higher for patients with high maximum NeoScore compared to lower maximum NeoScore. Overall, these results suggest an improved association of the NeoScore with survival following treatment with immunotherapy, compared with tumor mutational burden, in tumors with high tumor mutational burden.

## Discussion

Prioritization of immunogenic neoantigens is critical for applications to both the development of personalized vaccines and the identification of patients that are likely to benefit from treatment with immune checkpoint inhibition. However, many neoantigen characteristics have been suggested in the literature to date with no consensus on which characteristics impact whether the neoantigen generates a T cell response. Additionally, no model has demonstrated both the ability to score each neoantigen with high sensitivity and specificity and significant association with survival in response to immune checkpoint inhibition. The successes of this study are as follows: 1) identification of those neoantigen characteristics of greatest importance in determining neoantigen immunogenicity, 2) the combination of these characteristics into a single overall immunogenicity score, the NeoScore, with practical applications to personalized vaccine development, 3) integration of the NeoScore into a web application for easy use, and 4) demonstration of the clinical significance of the NeoScore in melanoma.

A model-based statistical prediction approach was used to select the characteristics of SNV and small indel-derived neoantigens that were most predictive of immunogenicity. The dissociation constant and binding stability of the neoantigen:MHC class I complex, and the expression were the combination of neoantigen characteristics best able to discriminate between immunogenic and non-immunogenic neoantigens. While the identification of these three characteristics is consistent with the recent findings of the TESLA consortium (Wells *et al*. 2020), it is important to note that the approaches taken by our group and the original group differed in several key ways. First, a completely agnostic approach was applied to the selection of characteristics, including all characteristics that have been suggested in the literature to date; whereas the TESLA consortium began with those shown to have statistical significance. Taking a completely agnostic approach ensured the greatest potential to select the combination of characteristics that maximized the separation of immunogenic and non-immunogenic neoantigens. Second, additional characteristics were included that were not considered by the TESLA consortium, including variant allele frequency and sequence similarity to the closest matched human peptide. Finally, these results were expanded by combining the characteristics into an overall immunogenicity score, the NeoScore. A web application has been made available to calculate the NeoScore or the abbreviated NeoScore and provide a list of neoantigens prioritized by their predicted immunogenicity. The web application is expected to streamline the application of NeoScore for research purposes.

One of the key advantages of the NeoScore is that it allows for the prioritization of neoantigens that may not exceed the single set of thresholds provided by the TESLA consortium. While a threshold for the NeoScore was optimized in the TESLA dataset and demonstrated strong performance across the test datasets, the optimized threshold is necessarily conservative for two reasons. First, all datasets used to build and test the NeoScore consisted of neoantigens that had already been prioritized by the original group. Thus, the NeoScore is trained to discriminate between top candidates. Second, the NeoScore is trained on the natural T cell responses to neoantigens with no stimulation by a therapeutic agent. Treatment with immunotherapy or personalized vaccines may be able to elicit an immune response to neoantigens with a lower NeoScore. The conservativeness of the optimized threshold is highlighted by the fact that lower thresholds are selected for survival analysis, which suggests that a patient with a maximum NeoScore below the threshold may still be able to elicit an immune response to the tumor in the context of immunotherapy. Therefore, all neoantigens can be ranked in order of their NeoScore. Researchers applying NeoScore to a new dataset can first rank the predicted neoantigens, then decide on the optimal number of neoantigens to test for a patient based on the unique neoantigen profile of the given patient.

A surprising finding is the lack of association between mutational burden and progression-free survival in the Van Allen dataset and the association of increased mutational burden with decreased progression-free survival in the Liu dataset. These results are inconsistent with the association of increased mutation burden with increased response rate to immune checkpoint inhibition observed across cancer types (Yarchoan, Hopkins, and Jaffee 2017). An explanation for the lack of association of mutation burden with survival following treatment with immune checkpoint inhibition is that the cutaneous melanoma subset consists of all tumors with a high mutational burden. A tumor with a particularly low mutational burden may not have any neoantigens, causing it to have a poor response to immune checkpoint inhibition. However, a high mutational burden alone does not guarantee a good response to immune checkpoint inhibition. For example, the patient in the Liu dataset with the lowest progression-free survival had 12,973 mutations, but not a single neoantigen predicted to be immunogenic. Similarly, the patient with the second lowest progression-free survival in the Van Allen dataset had 22,666 immunogenic neoantigens with none predicted to be immunogenic. Both cases highlight that, for tumors with a high mutational burden, there are additional characteristics beyond mutational burden that are of importance in determining the immune response. As demonstrated by our work, a high maximum NeoScore has an improved association with progression-free survival compared to mutational burden.

The literature to date supports that mutational burden is associated with response to therapy across cancers (Yarchoan, Hopkins, and Jaffee 2017) or across cancer sub-types, but not within tumors with a high mutational burden. Within non-small cell lung cancer, there was a significant increase in response rates and survival in patients with a higher mutational burden than those with a lower mutational burden (Rizvi, Hellmann, *et al*. 2015). These results reflect a split between the patients with a mutational signature from smoking carcinogens and those with no evidence of exposure to smoking carcinogens. In contrast, within small cell lung cancers, which are nearly universally associated with smoking, there was a weaker association of mutational burden with response to therapy (Hellmann *et al*. 2019). Three independent studies demonstrated a significant association between high mutational burden and improved response to immune checkpoint inhibition with either anti-CTLA-4 or anti-PD-1 monoclonal antibody treatment in melanoma patients (Snyder *et al*. 2014; Van Allen *et al*. 2015, Liu *et al*. 2019). However, these studies included cutaneous and occult melanoma, which have a high mutational burden, as well as acral and mucosal melanoma, which have lower mutational burdens. As demonstrated here, there is no association between high mutational burden and improved progression-free survival in the Van Allen and Liu datasets when restricted to cutaneous melanoma. As noted by Snyder *et al*., the patient in their dataset with the highest number of mutations had minimal or no benefit from anti-CTLA-4 monoclonal antibody treatment. Overall, in combination with prior studies, our results highlight the importance of considering the immunogenicity of the neoantigen in predicting the response to immune checkpoint inhibition.

Despite the successes of our work to date, there are several limitations that highlight areas for future research. First, the maximum immunogenicity score is not able to account for the response to therapy of all patients. One possible reason that the NeoScore is not able to fully predict treatment response is that the model did not include neoantigens derived from gene fusions, products of noncanonical open reading frames, canonical open reading frames with a frameshift, or large indels. To our knowledge, there is no available dataset that has validated neoantigens derived from these sources. Given that recent work has demonstrated that a single neoantigen from a gene fusion product can drive complete tumor regression (Yang *et al*. 2019) and non-canonical proteins disproportionately generate MHC class I binding neoantigens (Ruiz Cuevas *et al*. 2021), patients misclassified by our model may have had neoantigens from one of these classes of mutations. Additionally, since each of the validated datasets tested the immunogenicity of neoantigens that had already been prioritized by the original group, there may be classes of neoantigens that are immunogenic but were not included in any test dataset. This is particularly important given recent evidence that there are classes of neoantigens that have very low binding to MHC class I (Brennick *et al*. 2020). The inclusion of neoantigens derived from a broader set of mutations may alter the neoantigen characteristics selected as important by our regularized regression approach. For example, neoantigens derived from gene fusions, large indels, or frameshifts are likely to have less sequence similarity to normal human peptides than an SNV or small indel-derived neoantigen, causing characteristics such as the sequence similarity or amplitude to be of greater importance. The need to consider additional types of mutations underscores the importance of generating a validated dataset, including these additional mutations, and repeating characteristic selection. A second reason that the NeoScore may not be able to account for the response to therapy of all patients is that the model does not consider whether the neoantigens that ranked as immunogenic were able to elicit a CD4+ T cell response. Studies have observed that the most effective vaccines are those with neoantigens that elicit a combined CD4+ and CD8+ response (Kreiter *et al*. 2015; Alspach *et al*. 2019; Ossendorp *et al*. 1998; Bennett *et al*. 1997; Xu, Kallinteris, and von Hofe 2012). These studies highlight the importance of creating and integrating a similar prioritization model for MHC class II-restricted neoantigens.

This work has successfully identified the key neoantigen characteristics associated with neoantigen immunogenicity using an agnostic approach that considered all the characteristics suggested by the literature to date. These characteristics have been integrated into a single, overall score, the NeoScore, that predicts the immunogenicity of each neoantigen with high sensitivity and specificity. Finally, the maximum NeoScore has improved association with progression-free survival in response to treatment with immune checkpoint inhibition in cutaneous melanoma compared to mutational burden. NeoScore is anticipated to improve neoantigen prioritization for the development of personalized vaccines and the determination of which patients are likely to respond to immunotherapy.

## Data Availability

WES and RNAseq data for the TESLA consortium dataset are available through Synapse. Validation data for the TESLA consortium data is available through the supplementary material from the Wells et al. manuscript. All tested neoantigens, expression data, and validation information for the test datasets are provided through supplementary data tables from their respective publications. RNAseq data for the Cohen dataset was obtained through the NCBI with accession number SRP062169. WES and RNAseq data for both the Liu and Van Allen datasets are accessible through dbgap with the accession number phs000452. Links to all datasets are provided below.

https://www.synapse.org/#!Synapse:syn21048999/wiki/603788

https://ars.els-cdn.com/content/image/1-s2.0-S0092867420311569-mmc4.xlsx

https://trace.ddbj.nig.ac.jp/DRASearch/study?acc=SRP062169

https://www.ncbi.nlm.nih.gov/pmc/articles/PMC4607110/bin/JCI82416sdt2.xls

https://science.sciencemag.org/highwire/filestream/679287/field_highwire_adjunct_files/2/aaf2288-Table-S8.xlsx

https://www.ncbi.nlm.nih.gov/pmc/articles/PMC4549796/bin/NIHMS713231-supplement-Tables_S1-S3.xlsx

https://www.ncbi.nlm.nih.gov/pmc/articles/PMC5577644/bin/NIHMS892660-supplement-Supp_4.pdf

https://www.ncbi.nlm.nih.gov/projects/gap/cgi-bin/study.cgi?study_id=phs000452.v3.p1

## Acknowledgments

We thank Dr. Tanya N. Phung for her advice and assistance in comparing the software for the identification of SNVs and indels and integrating the pipeline from raw whole-exome sequencing data to potential neoantigens. We thank Dr. Chi Zhou for his assistance with the pTuneos software. We thank Anngela Adams for her assistance in polishing figures. This work was supported in part by the Springboard Initiative from the University of Arizona College of Medicine-Phoenix (K.T.H), and the University of Arizona College of Medicine-Phoenix M.D./Ph.D. Program (E.S.B).

## Author contributions

Conceptualization E.S.B., K.H.B., M.A.W., and K.T.H. Methodology E.S.B, B.J.L., M.A.W., and K.T.H. Software E.S.B. Formal Analysis E.S.B. and B.J.L. Investigation E.S.B. Resources K.H.B., M.A.W., and K.T.H. Writing -- Original Draft E.S.B. and K.T.H. Writing -- Reviewing and Editing E.S.B., K.H.B., B.J.L., M.A.W., and K.T.H. Visualization E.S.B. Supervision K.T.H. Funding Acquisition K.T.H.

## Methods

### Training datasets

For training, whole-exome sequencing (WES) and RNA sequencing (RNAseq) data were obtained from the TESLA consortium database on Synapse (Wells *et al*. 2020). The data come from four patients with melanoma and a single lung cancer patient. Neoantigens were used for model creation if they were 1) tested for immunogenicity by the TESLA consortium, 2) identified by either GATK Mutect2 or Strelka, and 3) had expression data. A description of the datasets used, and all relevant accession details are provided in Supplementary Table 2.

### Test datasets

For testing of the model, lists of tested neoantigens from melanoma tumors were obtained (Strønen *et al*. 2016; Cohen *et al*. 2015; Carreno *et al*. 2015; Ott *et al*. 2017). These datasets contain 357 tested neoantigens (n=7 immunogenic neoantigens) (Cohen *et al*. 2015), 149 tested neoantigens (n=18 immunogenic neoantigens) (Ott *et al*. 2017), 57 tested neoantigens (n=11 immunogenic neoantigens) (Strønen *et al*. 2016) and 21 tested neoantigens (n=9 immunogenic neoantigens) (Carreno *et al*. 2015). The neoantigens were tested for immunogenicity with tetramer staining and cytokine secretion (Cohen *et al*. 2015), ELISPOT (Ott *et al*. 2017), and multimer staining (Carreno *et al*. 2015; Strønen *et al*. 2016). Cohen and Carreno also provided the expression data quantified as either fragments per kilobase of transcript per million mapped reads (FPKM), reads per kilobase of transcript per million mapped reads (RPKM), or transcripts per million (TPM) (Strønen *et al*. 2016; Cohen *et al*. 2015; Carreno *et al*. 2015; Ott *et al*. 2017). For the Cohen dataset, no expression data was provided. The RNAseq data from the Cohen dataset was obtained and analyzed as described below for read count quantification. Accession information is in Supplementary Table 2.

### Survival analysis datasets

For survival analysis, WES and RNAseq data were obtained from the Van Allen *et al*. and Liu *et al*. cohort (all accession information is in Supplementary Table 2). The inclusion criteria for the Van Allen dataset were as follows: 1) both WES and RNAseq data available, 2) cutaneous melanoma as the primary lesion. All patients in the Van Allen cohort underwent treatment with an anti-CTLA-4 monoclonal antibody (ipilimumab). Inclusion criteria for the Liu *et al* dataset were as follows: 1) both WES and RNAseq data available, 2) cutaneous melanoma as the primary lesion, 3) no prior treatment with an anti-CTLA-4 monoclonal antibody, and 4) HLA types identifiable with arcasHLA (Orenbuch *et al*. 2020). All patients in the Liu cohort underwent treatment with an anti-PD-1 monoclonal antibody (nivolumab or pembrolizumab).

### Quality control and trimming

WES and RNAseq FASTQ files from the TESLA consortium and Liu *et al*. and RNAseq FASTQ files from Cohen *et al*. were visualized for quality using FastQC (https://www.bioinformatics.babraham.ac.uk/projects/fastqc/). FASTQ files were trimmed for quality using Trimmomatic (Bolger, Lohse, and Usadel 2014) IlluminaClip with the following parameters: seed_mismatches = 2, palindrome_clip_threshold = 30, simple_clip_threshold = 10, leading = 10, trailing = 10, winsize = 4, winqual = 15. Quality was then re-visualized after trimming.

### Read mapping

Trimmed WES reads were mapped to the GRCh38.p7 reference genome, from 1000 genomes (Clarke *et al*. 2017), and read group labels were added using BWA-mem (Li 2013). SAM files were converted to BAM, coordinate sorted, and read mates were corrected using SAMtools v. 1.4 (Li *et al*. 2009). The BAM files were then converted to pileup format using SAMtools v. 1.4 (Li *et al*. 2009).

### Identification of somatic mutations

Single nucleotide variants (SNVs) and small insertions/deletions (indels) were identified using four programs, along with their recommended filters: GATK Mutect2 version 4.1.7.0 with default parameters, Varscan2 version 2.3.9 with minimum coverage of 10, minimum variant allele frequency of 0.08, and somatic p-value of 0.05, Strelka version 2.9.2 with default parameters, and LoFreq version 2.1.1 with default parameters (Benjamin *et al*. 2019; Koboldt *et al*. 2012; Saunders *et al*. 2012; Wilm *et al*. 2012). Results for GATK Mutect2 were filtered with the recommended FilterMutectCalls, and Varscan2 results were filtered using the Perl false-positive filter (https://github.com/ckandoth/variant-filter). Results from all four programs were examined with and without their respective filters. LoFreq results were not filtered to allow maximal potential to identify the missing mutations. Matched normal samples were used as the reference for each sample.

### Variant annotation and neoantigen generation

Somatic mutations were separated from those SNVs that fell within 1 bp of an indel position, as these are likely to be false positives due to alignment errors. The mutations were annotated using the Variant Effect Predictor (VEP) tool from Ensembl version 90.9 (McLaren *et al*. 2016). Then, 17mer amino acid sequences were generated for each mutation using pVAC-Seq tools version 3.0.5 (Hundal *et al*. 2016). Finally, the 17mers were split into all possible 9 or 10mers where the mutation of interest was in every location. The full pipeline from read mapping through to the identification of somatic mutations is available at https://github.com/SexChrLab/Cancer_Genomics.

### Calculation of neoantigen characteristics

For each of the validated neoantigens, 10 separate neoantigen characteristics with potential significance in predicting expression, processing, MHC binding, and T cell recognition potential were calculated as described here. The full pipeline for calculation and processing of the neoantigen characteristics can be found at https://github.com/ElizabethBorden/Process_peptide_lists. A log transformation was applied if the distribution of the characteristic had a large degree of skewness. The difference in the values for each of the neoantigen characteristics between immunogenic and non-immunogenic neoantigens was assessed using a two-sample, two-sided t-test. Correlation coefficients were calculated using Spearman correlation coefficients. P-values below 0.05 were considered statistically significant.

### Expression

For the datasets from Cohen *et al*., the TESLA consortium, and Liu *et al*., transcriptome assembly and read count quantifications were completed with Salmon version 0.11.3, using the Ensembl GRCh38.p7 reference genome (Cunningham *et al*. 2015; Patro *et al*. 2017). mRNA expression in units of TPM was log-transformed, and a constant of 0.1 was added to all values before the transformation to avoid taking the log of zero. To account for the different units used across each dataset, expression values were centered and normalized by subtracting the mean and then dividing by the standard deviation.

### Clonality

Copy number variation was calculated with sequenza (Favero *et al*. 2015), and clonality was calculated using the deconvolution software, FastClone (Xiao *et al*. 2020).

### Variant allele frequency

Variant allele frequency (VAF) as calculated by GATK Mutect2 (Benjamin *et al*. 2019). Since the VAF was only calculated by GATK Mutect2, but not for Strelka, 14 missing data points were estimated as the average of the rest of the data. No statistically significant difference in the VAF was observed between immunogenic and non-immunogenic neoantigens, either with or without these data points.

### Processing

Cleavage and TAP transport potentials were calculated for each of the available neoantigens using NetCTLpan1.0 (Stranzl *et al*. 2010).

### Dissociation constants

Dissociation constants of the neoantigen:MHC class I complex were calculated in nanomolar (nM) units using NetMHCpan4.0 (Reynisson *et al*. 2020). These values were log-transformed before inclusion in the model.

### Binding stability

Binding stability of the neoantigen:MHC class I complex was then calculated as the half-life in units of hours using NetMHCstabpan1.0 (Reynisson *et al*. 2020; Rasmussen *et al*. 2016). These values were log-transformed before inclusion in the model.

### Hydrophobicity method from the TESLA consortium

The number of hydrophobic residues was divided by the total number of residues in the neoantigen to create a “hydrophobicity fraction” (Wells *et al*. 2020). In keeping with the methods of the TESLA consortium, hydrophobic residues here were isoleucine, leucine, phenylalanine, methionine, tryptophan, valine, and cysteine.

### Hydrophobicity with empirical prevalence

The hydrophobicity fraction was calculated as described for the TESLA consortium using the amino acids found to be empirically prevalent (Chowell *et al*. 2015). Proline, leucine, and methionine were considered to have a high probability and given a score of +2. Glycine, tryptophan, phenylalanine, isoleucine, and valine were found to have a medium probability and given a score of +1. All others were given a score of 0.

### Hydrophobicity Łuksza method

A neoantigen was given a score of zero if the mutation at the anchor residue changed from a hydrophobic to a hydrophilic residue (Łuksza *et al*. 2017). All other neoantigens were given a score of one. Hydrophobic neoantigens here were defined as alanine, isoleucine, leucine, methionine, phenylalanine, tryptophan, tyrosine, and valine.

### TCR recognition

Prediction of TCR recognition potential, R, was calculated as described (Łuksza *et al*. 2017). A BLOSUM62 similarity matrix was used to assess the sequence similarity between a neoantigen and the closest matched known T cell epitope from the Immune Epitope Database (IEDB) (Vita *et al*. 2015). The sequence similarity was then used in place of binding energies, and the TCR recognition potential was calculated as:

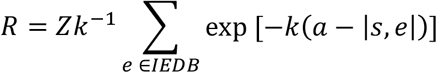

Where |*s, e*| is the sequence similarity, *a* is the horizontal displacement of the binding curve, and *k* sets the steepness of the curve at *a*. Based on the model fit by Łuksza *et al*., the parameters *a* and *k* were set to 26 and 4.87 respectively (Łuksza *et al*. 2017). These parameters were optimized for both melanoma and lung cancer patients, the two cancer populations included in our training and test datasets. Finally, Z(k) is the partition function over the unbound state and all bound states, calculated as follows:

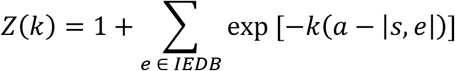

### Sequence Similarity

The closest matched human peptide was identified using Blast v. 2.10.1 (Camacho *et al*. 2009), and the sequence similarity of the potential neoantigen to the closest matched human peptide was calculated using a BLOSUM62 matrix, as described (Wood *et al*. 2018). The sequence similarity was normalized across neoantigen length by dividing by the number of amino acids.

### Amplitude

Dissociation constants for the neoantigen:MHC class I complex were then adjusted by the dissociation constants for the closest matched normal human peptide:MHC class I complex, a characteristic called amplitude. The ratio was taken of the dissociation constant of the closest matched human peptide:MHC class I 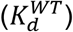 to the dissociation constant of the neoantigen:MHC class I 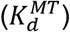 as follows (Łuksza *et al*. 2017):

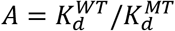

### Analysis of anchor vs. non-anchor residue mutations

Neoantigens were separated by their mutation position (anchor vs. non-anchor residues). Then, both the amplitude and the dissociation constant of the neoantigen:MHC class I complex were compared between the immunogenic and non-immunogenic neoantigens within each group. Comparisons were done using a two-sample, two-sided t-test. P-values below 0.05 were considered statistically significant.

### HLA typing

HLA types were identified on normal tissue samples for each patient in the Liu *et al*. dataset using arcasHLA (Orenbuch *et al*. 2020). One limitation of the arcasHLA method is that all 6 HLA alleles were not determined for each patient and at least one HLA (A, B, or C) was excluded for 18 patients. HLA types provided in the supplementary data were used for each patient in the Van Allen *et al*. dataset (Van Allen *et al*. 2015).

### Elastic net regression modeling

The TESLA consortium data was split into 1000 random subgroups containing all the immunogenic neoantigens (n=26) and an equal number of randomly selected non-immunogenic neoantigens. An elastic net regression was performed using the glmnet package in R for each of these splits (Engebretsen and Bohlin 2019). The selected coefficients and area under the receiver operator characteristics curve (AUC) from each model were tabulated across the 1000 splits.

### Logistic regression modeling

Logistic regression modeling was performed with the RMS package in R statistical software (https://cran.r-project.org/web/packages/rms/rms.pdf).

### Survival analysis

To avoid scaling expression values on a patient-level basis, coefficients were determined on the TESLA consortium data for log-transformed, non-scaled TPM expression data. The intercept changed to -4.0494 and the expression coefficient to 0.4749, but there was no change the coefficients of the dissociation constant or binding stability and no affect the performance of the model. The optimal threshold for each of the mutational burden, neoantigen burden, and maximum NeoScore were determined using MaxStat (Hothorn 2007). Then the survival analysis was performed between scores above the selected threshold and below using a Kaplan Meier analysis and a log-rank test for statistical significance. P-values below 0.05 were considered statistically significant.

## References

Alspach, E., Lussier, D.M., Miceli, A.P., Kizhvatov, I., DuPage, M., Luoma, A.M., Meng, W., Lichti, C.F., Esaulova, E., Vomund, A.N., et al. (2019). MHC-II neoantigens shape tumour immunity and response to immunotherapy. Nature 574, 696–701. https://doi.org/10.1038/s41586-019-1671-8

Anagnostou, V., Smith, K.N., Forde, P.M., Niknafs, N., Bhattacharya, R., White, J., Zhang, T., Adleff, V., Phallen, J., Wali, N., et al. (2017). Evolution of Neoantigen Landscape during Immune Checkpoint Blockade in Non-Small Cell Lung Cancer. Cancer Discov. 7, 264–276. https://doi.org/10.1158/2159-8290.cd-16-0828

Benjamin, D., Sato, T., Cibulskis, K., Getz, G., Stewart, C., and Lichtenstein, L. (2019). Calling Somatic SNVs and Indels with Mutect2. https://doi.org/10.1101/861054

Bennett, S.R., Carbone, F.R., Karamalis, F., Miller, J.F., and Heath, W.R. (1997). Induction of a CD8+ cytotoxic T lymphocyte response by cross-priming requires cognate CD4+ T cell help. J. Exp. Med. 186, 65–70. https://dx.doi.org/10.1084%2Fjem.186.1.65

Bjerregaard, A.-M., Nielsen, M., Hadrup, S.R., Szallasi, Z., and Eklund, A.C. (2017). MuPeXI: prediction of neo-epitopes from tumor sequencing data. Cancer Immunol. Immunother. 66, 1123–1130. https://doi.org/10.1007/s00262-017-2001-3

Bolger, A.M., Lohse, M., and Usadel, B. (2014). Trimmomatic: a flexible trimmer for Illumina sequence data. Bioinformatics 30, 2114–2120. https://doi.org/10.1093/bioinformatics/btu170

Brennick, C.A., George, M.M., Srivastava, P.K., and Karandikar, S.H. (2020). Prediction of cancer neoepitopes needs new rules. Semin. Immunol. 47, 101387. https://doi.org/10.1016/j.smim.2020.101387

Camacho, C., Coulouris, G., Avagyan, V., Ma, N., Papadopoulos, J., Bealer, K., and Madden, T.L. (2009). BLAST+: architecture and applications. BMC Bioinformatics 10, 1–9. https://doi.org/10.1186/1471-2105-10-421

Capietto, A.-H., Jhunjhunwala, S., Pollock, S.B., Lupardus, P., Wong, J., Hänsch, L., Cevallos, J., Chestnut, Y., Fernandez, A., Lounsbury, N., et al. (2020). Mutation position is an important determinant for predicting cancer neoantigens. J. Exp. Med. 217. https://doi.org/10.1084/jem.20190179

Carreno, B.M., Magrini, V., Becker-Hapak, M., Kaabinejadian, S., Hundal, J., Petti, A.A., Ly, A., Lie, W.-R., Hildebrand, W.H., Mardis, E.R., et al. (2015). Cancer immunotherapy. A dendritic cell vaccine increases the breadth and diversity of melanoma neoantigen-specific T cells. Science 348, 803–808. https://doi.org/10.1126/science.aaa3828

Chowell, D., Krishna, S., Becker, P.D., Cocita, C., Shu, J., Tan, X., Greenberg, P.D., Klavinskis, L.S., Blattman, J.N., and Anderson, K.S. (2015). TCR contact residue hydrophobicity is a hallmark of immunogenic CD8+ T cell epitopes. Proc. Natl. Acad. Sci. U. S. A. 112, E1754–E1762. https://doi.org/10.1073/pnas.1500973112

Clarke, L., Fairley, S., Zheng-Bradley, X., Streeter, I., Perry, E., Lowy, E., Tassé, A.-M., and Flicek, P. (2017). The international Genome sample resource (IGSR): A worldwide collection of genome variation incorporating the 1000 Genomes Project data. Nucleic Acids Res. 45, D854–D859. https://dx.doi.org/10.1093%2Fnar%2Fgkz836

Cohen, C.J., Gartner, J.J., Horovitz-Fried, M., Shamalov, K., Trebska-McGowan, K., Bliskovsky, V.V., Parkhurst, M.R., Ankri, C., Prickett, T.D., Crystal, J.S., et al. (2015). Isolation of neoantigen-specific T cells from tumor and peripheral lymphocytes. J. Clin. Invest. 125, 3981–3991. https://doi.org/10.1172/jci82416

Cunningham, F., Amode, M.R., Barrell, D., Beal, K., Billis, K., Brent, S., Carvalho-Silva, D., Clapham, P., Coates, G., Fitzgerald, S., et al. (2015). Ensembl 2015. Nucleic Acids Res. 43, D662–D669. https://doi.org/10.1093/nar/gku1010

Engebretsen, S., and Bohlin, J. (2019). Statistical predictions with glmnet. Clin. Epigenetics 11, 123. https://doi.org/10.1186/s13148-019-0730-1

Favero, F., Joshi, T., Marquard, A.M., Birkbak, N.J., Krzystanek, M., Li, Q., Szallasi, Z., and Eklund, A.C. (2015). Sequenza: allele-specific copy number and mutation profiles from tumor sequencing data. Ann. Oncol. 26, 64–70. https://doi.org/10.1093/annonc/mdu479

Gubin, M.M., Zhang, X., Schuster, H., Caron, E., Ward, J.P., Noguchi, T., Ivanova, Y., Hundal, J., Arthur, C.D., Krebber, W.-J., et al. (2014). Checkpoint blockade cancer immunotherapy targets tumour-specific mutant antigens. Nature 515, 577–581. https://doi.org/10.1038/nature13988

Gubin, M.M., Artyomov, M.N., Mardis, E.R., and Schreiber, R.D. (2015). Tumor neoantigens: building a framework for personalized cancer immunotherapy. J. Clin. Invest. 125, 3413–3421. https://doi.org/10.1172/jci80008

Hellmann, M.D., Callahan, M.K., Awad, M.M., Calvo, E., Ascierto, P.A., Atmaca, A., Rizvi, N.A., Hirsch, F.R., Selvaggi, G., Szustakowski, J.D., et al. (2019). Tumor Mutational Burden and Efficacy of Nivolumab Monotherapy and in Combination with Ipilimumab in Small-Cell Lung Cancer. Cancer Cell 35, 329. https://doi.org/10.1016/j.ccell.2018.04.001

Hilf, N., Kuttruff-Coqui, S., Frenzel, K., Bukur, V., Stevanović, S., Gouttefangeas, C., Platten, M., Tabatabai, G., Dutoit, V., van der Burg, S.H., et al. (2019). Actively personalized vaccination trial for newly diagnosed glioblastoma. Nature 565, 240–245. https://doi.org/10.1038/s41586-018-0810-y

Hodi, F.S., Chiarion-Sileni, V., Gonzalez, R., Grob, J.-J., Rutkowski, P., Cowey, C.L., Lao, C.D., Schadendorf, D., Wagstaff, J., Dummer, R., et al. (2018). Nivolumab plus ipilimumab or nivolumab alone versus ipilimumab alone in advanced melanoma (CheckMate 067): 4-year outcomes of a multicentre, randomised, phase 3 trial. Lancet Oncol. 19, 1480–1492. https://doi.org/10.1016/s1470-2045(18)30700-9

Hosmer, D.W., Jr, Lemeshow, S., and Sturdivant, R.X. (2013). Applied Logistic Regression (John Wiley & Sons). https://doi.org/10.1002/9781118548387

Hothorn, T. (2007). Maxstat: maximally selected rank statistics. https://cran.r-project.org/web/packages/maxstat/vignettes/maxstat.pdf

Hundal, J., Carreno, B.M., Petti, A.A., Linette, G.P., Griffith, O.L., Mardis, E.R., and Griffith, M. (2016). pVAC-Seq: A genome-guided in silico approach to identifying tumor neoantigens. Genome Med. 8, 11. https://doi.org/10.1186/s13073-016-0264-5

Keskin, D.B., Anandappa, A.J., Sun, J., Tirosh, I., Mathewson, N.D., Li, S., Oliveira, G., Giobbie-Hurder, A., Felt, K., Gjini, E., et al. (2019). Neoantigen vaccine generates intratumoral T cell responses in phase Ib glioblastoma trial. Nature 565, 234–239. https://doi.org/10.1038/s41586-018-0792-9

Kim, S., Kim, H.S., Kim, E., Lee, M.G., Shin, E.-C., Paik, S., and Kim, S. (2018). Neopepsee: accurate genome-level prediction of neoantigens by harnessing sequence and amino acid immunogenicity information. Ann. Oncol. 29, 1030–1036. https://doi.org/10.1093/annonc/mdy022

Koboldt, D.C., Zhang, Q., Larson, D.E., Shen, D., McLellan, M.D., Lin, L., Miller, C.A., Mardis, E.R., Ding, L., and Wilson, R.K. (2012). VarScan 2: somatic mutation and copy number alteration discovery in cancer by exome sequencing. Genome Res. 22, 568–576. https://doi.org/10.1101/gr.129684.111

Kreiter, S., Vormehr, M., van de Roemer, N., Diken, M., Löwer, M., Diekmann, J., Boegel, S., Schrörs, B., Vascotto, F., Castle, J.C., et al. (2015). Erratum: Mutant MHC class II epitopes drive therapeutic immune responses to cancer. Nature 523, 370. https://doi.org/10.1038/nature14426

Li, H. (2013). Aligning sequence reads, clone sequences and assembly contigs with BWA-MEM. https://arxiv.org/pdf/1303.3997.pdf

Li, H., Handsaker, B., Wysoker, A., Fennell, T., Ruan, J., Homer, N., Marth, G., Abecasis, G., Durbin, R., and 1000 Genome Project Data Processing Subgroup (2009). The Sequence Alignment/Map format and SAMtools. Bioinformatics 25, 2078–2079. https://doi.org/10.1093/bioinformatics/btp352

Linette, G.P., Becker-Hapak, M., Skidmore, Z.L., Baroja, M.L., Xu, C., Hundal, J., Spencer, D.H., Fu, W., Cummins, C., Robnett, M., et al. (2019). Immunological ignorance is an enabling feature of the oligo-clonal T cell response to melanoma neoantigens. Proc. Natl. Acad. Sci. U. S. A. 116, 23662–23670. https://doi.org/10.1073/pnas.1906026116

Liu, D., Schilling, B., Liu, D., Sucker, A., Livingstone, E., Jerby-Arnon, L., Zimmer, L., Gutzmer, R., Satzger, I., Loquai, C., et al. (2019). Integrative molecular and clinical modeling of clinical outcomes to PD1 blockade in patients with metastatic melanoma. Nat. Med. 25, 1916–1927. https://doi.org/10.1038/s41591-019-0654-5

Lommatzsch, M., Bratke, K., and Stoll, P. (2018). Neoadjuvant PD-1 Blockade in Resectable Lung Cancer. N. Engl. J. Med. 379, e14. https://doi.org/10.1056/nejmc1808251

Łuksza, M., Riaz, N., Makarov, V., Balachandran, V.P., Hellmann, M.D., Solovyov, A., Rizvi, N.A., Merghoub, T., Levine, A.J., Chan, T.A., et al. (2017). A neoantigen fitness model predicts tumour response to checkpoint blockade immunotherapy. Nature 551, 517–520. https://doi.org/10.1038/nature24473

McGranahan, N., Furness, A.J.S., Rosenthal, R., Ramskov, S., Lyngaa, R., Saini, S.K., Jamal-Hanjani, M., Wilson, G.A., Birkbak, N.J., Hiley, C.T., et al. (2016). Clonal neoantigens elicit T cell immunoreactivity and sensitivity to immune checkpoint blockade. Science 351, 1463–1469. https://doi.org/10.1126/science.aaf1490

McLaren, W., Gil, L., Hunt, S.E., Riat, H.S., Ritchie, G.R.S., Thormann, A., Flicek, P., and Cunningham, F. (2016). The Ensembl Variant Effect Predictor. Genome Biol. 17, 122. https://doi.org/10.1186/s13059-016-0974-4

Miller, A., Asmann, Y., Cattaneo, L., Braggio, E., Keats, J., Auclair, D., Lonial, S., MMRF CoMMpass Network, Russell, S.J., and Stewart, A.K. (2017). High somatic mutation and neoantigen burden are correlated with decreased progression-free survival in multiple myeloma. Blood Cancer J. 7, e612. https://doi.org/10.1038/bcj.2017.94

Mittal, D., Gubin, M.M., Schreiber, R.D., and Smyth, M.J. (2014). New insights into cancer immunoediting and its three component phases--elimination, equilibrium and escape. Curr. Opin. Immunol. 27, 16–25. https://doi.org/10.1016/j.coi.2014.01.004

Orenbuch, R., Filip, I., Comito, D., Shaman, J., Pe’er, I., and Rabadan, R. (2020). arcasHLA: high-resolution HLA typing from RNAseq. Bioinformatics 36, 33–40. https://doi.org/10.1093/bioinformatics/btz474

Ossendorp, F., Mengedé, E., Camps, M., Filius, R., and Melief, C.J. (1998). Specific T helper cell requirement for optimal induction of cytotoxic T lymphocytes against major histocompatibility complex class II negative tumors. J. Exp. Med. 187, 693–702. https://doi.org/10.1084/jem.187.5.693

Ott, P.A., Hu, Z., Keskin, D.B., Shukla, S.A., Sun, J., Bozym, D.J., Zhang, W., Luoma, A., Giobbie-Hurder, A., Peter, L., et al. (2017). An immunogenic personal neoantigen vaccine for patients with melanoma. Nature 547, 217–221. https://doi.org/10.1038/nature22991

Patro, R., Duggal, G., Love, M.I., Irizarry, R.A., and Kingsford, C. (2017). Salmon provides fast and bias-aware quantification of transcript expression. Nat. Methods 14, 417–419. https://doi.org/10.1038/nmeth.4197

Rasmussen, M., Fenoy, E., Harndahl, M., Kristensen, A.B., Nielsen, I.K., Nielsen, M., and Buus, S. (2016). Pan-Specific Prediction of Peptide-MHC Class I Complex Stability, a Correlate of T Cell Immunogenicity. J. Immunol. 197, 1517–1524. https://doi.org/10.4049/jimmunol.1600582

Rausch, M.P., and Hastings, K.T. (2017). Immune Checkpoint Inhibitors in the Treatment of Melanoma: From Basic Science to Clinical Application. In Cutaneous Melanoma: Etiology and Therapy, W.H. Ward, and J.M. Farma, eds. (Brisbane (AU): Codon Publications). https://doi.org/10.15586/codon.cutaneousmelanoma.2017.ch9

Reynisson, B., Alvarez, B., Paul, S., Peters, B., and Nielsen, M. (2020). NetMHCpan-4.1 and NetMHCIIpan-4.0: improved predictions of MHC antigen presentation by concurrent motif deconvolution and integration of MS MHC eluted ligand data. Nucleic Acids Res. 48, W449–W454. https://doi.org/10.1093/nar/gkaa379

Rizvi, N.A., Hellmann, M.D., Snyder, A., Kvistborg, P., Makarov, V., Havel, J.J., Lee, W., Yuan, J., Wong, P., Ho, T.S., et al. (2015a). Cancer immunology. Mutational landscape determines sensitivity to PD-1 blockade in non-small cell lung cancer. Science 348, 124–128. https://doi.org/10.1126/science.aaa1348

Rizvi, N.A., Garon, E.B., Leighl, N., Hellmann, M.D., Patnaik, A., Gandhi, L., Eder, J.P., Rangwala, R.A., Lubiniecki, G., Zhang, J., et al. (2015b). Optimizing PD-L1 as a biomarker of response with pembrolizumab (pembro; MK-3475) as first-line therapy for PD-L1–positive metastatic non-small cell lung cancer (NSCLC): Updated data from KEYNOTE-001. Journal of Clinical Oncology 33, 8026–8026. https://doi.org/10.1200/jco.2015.33.15_suppl.8026

Ruiz Cuevas, M.V., Hardy, M.-P., Hollý, J., Bonneil, É., Durette, C., Courcelles, M., Lanoix, J., Côté, C., Staudt, L.M., Lemieux, S., et al. (2021). Most non-canonical proteins uniquely populate the proteome or immunopeptidome. Cell Rep. 34, 108815. https://dx.doi.org/10.1016%2Fj.celrep.2021.108815

Sahin, U., Derhovanessian, E., Miller, M., Kloke, B.-P., Simon, P., Löwer, M., Bukur, V., Tadmor, A.D., Luxemburger, U., Schrörs, B., et al. (2017). Personalized RNA mutanome vaccines mobilize poly-specific therapeutic immunity against cancer. Nature 547, 222–226. https://doi.org/10.1038/nature23003

Saunders, C.T., Wong, W.S.W., Swamy, S., Becq, J., Murray, L.J., and Cheetham, R.K. (2012). Strelka: accurate somatic small-variant calling from sequenced tumor-normal sample pairs. Bioinformatics 28, 1811–1817. https://doi.org/10.1093/bioinformatics/bts271

Schmidt, J., Smith, A.R., Magnin, M., Racle, J., Devlin, J.R., Bobisse, S., Cesbron, J., Bonnet, V., Carmona, S.J., Huber, F., et al. (2021). Prediction of neo-epitope immunogenicity reveals TCR recognition determinants and provides insight into immunoediting. Cell Rep Med 2, 100194. https://doi.org/10.1016/j.xcrm.2021.100194

Schumacher, T.N., and Schreiber, R.D. (2015). Neoantigens in cancer immunotherapy. Science 348, 69–74. https://doi.org/10.1126/science.aaa4971

Shemesh, C.S., Hsu, J.C., Hosseini, I., Shen, B.-Q., Rotte, A., Twomey, P., Girish, S., and Wu, B. (2021). Personalized Cancer Vaccines: Clinical Landscape, Challenges, and Opportunities. Mol. Ther. 29, 555– 570. https://doi.org/10.1016/j.ymthe.2020.09.038

Snyder, A., Makarov, V., Merghoub, T., Yuan, J., Zaretsky, J.M., Desrichard, A., Walsh, L.A., Postow, M.A., Wong, P., Ho, T.S., et al. (2014). Genetic basis for clinical response to CTLA-4 blockade in melanoma. N. Engl. J. Med. 371, 2189–2199. https://doi.org/10.1056/nejmoa1406498

Solomon, B.J., Beavis, P.A., and Darcy, P.K. (2020). Promising Immuno-Oncology Options for the Future: Cellular Therapies and Personalized Cancer Vaccines. Am Soc Clin Oncol Educ Book 40, 1–6. https://doi.org/10.1200/edbk_281101

Stranzl, T., Larsen, M.V., Lundegaard, C., and Nielsen, M. (2010). NetCTLpan: pan-specific MHC class I pathway epitope predictions. Immunogenetics 62, 357–368. https://doi.org/10.1007/s00251-010-0441-4

Strønen, E., Toebes, M., Kelderman, S., van Buuren, M.M., Yang, W., van Rooij, N., Donia, M., Böschen, M.-L., Lund-Johansen, F., Olweus, J., et al. (2016). Targeting of cancer neoantigens with donor-derived T cell receptor repertoires. Science 352, 1337–1341. https://doi.org/10.1126/science.aaf2288

Tran, E., Turcotte, S., Gros, A., Robbins, P.F., Lu, Y.-C., Dudley, M.E., Wunderlich, J.R., Somerville, R.P., Hogan, K., Hinrichs, C.S., et al. (2014). Cancer Immunotherapy Based on Mutation-Specific CD4+ T Cells in a Patient with Epithelial Cancer. Science 344, 641–645. https://doi.org/10.1126/science.1251102

Tran, E., Robbins, P.F., and Rosenberg, S.A. (2017). “Final common pathway” of human cancer immunotherapy: targeting random somatic mutations. Nat. Immunol. 18, 255–262. https://doi.org/10.1038/ni.3682

Van Allen, E.M., Miao, D., Schilling, B., Shukla, S.A., Blank, C., Zimmer, L., Sucker, A., Hillen, U., Foppen, M.H.G., Goldinger, S.M., et al. (2015). Genomic correlates of response to CTLA-4 blockade in metastatic melanoma. Science 350, 207–211. https://doi.org/10.1126/science.aad0095

Vita, R., Overton, J.A., Greenbaum, J.A., Ponomarenko, J., Clark, J.D., Cantrell, J.R., Wheeler, D.K., Gabbard, J.L., Hix, D., Sette, A., et al. (2015). The immune epitope database (IEDB) 3.0. Nucleic Acids Res. 43. https://doi.org/10.1093/nar/gku938

Ward, J.P., Gubin, M.M., and Schreiber, R.D. (2016). The Role of Neoantigens in Naturally Occurring and Therapeutically Induced Immune Responses to Cancer. Adv. Immunol. 130, 25–74. https://doi.org/10.1016/bs.ai.2016.01.001

Wells, D.K., van Buuren, M.M., Dang, K.K., Hubbard-Lucey, V.M., Sheehan, K.C.F., Campbell, K.M., Lamb, A., Ward, J.P., Sidney, J., Blazquez, A.B., et al. (2020). Key Parameters of Tumor Epitope Immunogenicity Revealed Through a Consortium Approach Improve Neoantigen Prediction. Cell 183, 818–834.e13. https://doi.org/10.1016/j.cell.2020.09.015

Wilm, A., Aw, P.P.K., Bertrand, D., Yeo, G.H.T., Ong, S.H., Wong, C.H., Khor, C.C., Petric, R., Hibberd, M.L., and Nagarajan, N. (2012). LoFreq: a sequence-quality aware, ultra-sensitive variant caller for uncovering cell-population heterogeneity from high-throughput sequencing datasets. Nucleic Acids Res. 40, 11189– 11201. https://doi.org/10.1093/nar/gks918

Wood, M.A., Paralkar, M., Paralkar, M.P., Nguyen, A., Struck, A.J., Ellrott, K., Margolin, A., Nellore, A., and Thompson, R.F. (2018). Population-level distribution and putative immunogenicity of cancer neoepitopes. BMC Cancer 18, 414. https://doi.org/10.1186/s12885-018-4325-6

Xiao, Y., Wang, X., Zhang, H., Ulintz, P.J., Li, H., and Guan, Y. (2020). FastClone is a probabilistic tool for deconvoluting tumor heterogeneity in bulk-sequencing samples. Nat. Commun. 11, 4469. https://doi.org/10.1038/s41467-020-18169-2

Xu, M., Kallinteris, N.L., and von Hofe, E. (2012). CD4+ T-cell activation for immunotherapy of malignancies using Ii-Key/MHC class II epitope hybrid vaccines. Vaccine 30, 2805–2810. https://doi.org/10.1016/j.vaccine.2012.02.031

Yang, W., Lee, K.-W., Srivastava, R.M., Kuo, F., Krishna, C., Chowell, D., Makarov, V., Hoen, D., Dalin, M.G., Wexler, L., et al. (2019). Immunogenic neoantigens derived from gene fusions stimulate T cell responses. Nat. Med. 25, 767–775. https://doi.org/10.1038/s41591-019-0434-2

Yarchoan, M., Johnson, B.A., 3rd, Lutz, E.R., Laheru, D.A., and Jaffee, E.M. (2017a). Targeting neoantigens to augment antitumour immunity. Nat. Rev. Cancer 17, 569. https://doi.org/10.1038/nrc.2016.154

Yarchoan, M., Hopkins, A., and Jaffee, E.M. (2017b). Tumor Mutational Burden and Response Rate to PD-1 Inhibition. N. Engl. J. Med. 377, 2500–2501. https://doi.org/10.1056/nejmc1713444

Yee, C., Thompson, J.A., Byrd, D., Riddell, S.R., Roche, P., Celis, E., and Greenberg, P.D. (2002). Adoptive T cell therapy using antigen-specific CD8+ T cell clones for the treatment of patients with metastatic melanoma: in vivo persistence, migration, and antitumor effect of transferred T cells. Proc. Natl. Acad. Sci. U. S. A. 99, 16168–16173. https://doi.org/10.1073/pnas.242600099

Zacharakis, N., Chinnasamy, H., Black, M., Xu, H., Lu, Y.-C., Zheng, Z., Pasetto, A., Langhan, M., Shelton, T., Prickett, T., et al. (2018). Immune recognition of somatic mutations leading to complete durable regression in metastatic breast cancer. Nat. Med. 24, 724–730. https://doi.org/10.1038/s41591-018-0040-8

Zhou, C., Wei, Z., Zhang, Z., Zhang, B., Zhu, C., Chen, K., Chuai, G., Qu, S., Xie, L., Gao, Y., et al. (2019). pTuneos: prioritizing tumor neoantigens from next-generation sequencing data. Genome Med. 11, 67. https://doi.org/10.1186/s13073-019-0679-x

