## Supplemental figures for "NeoScore integrates characteristics of the neoantigen:MHC class I interaction and expression to accurately prioritize immunogenic neoantigens"

***Supplementary Table 1.*** *Summary of the number of validated immunogenic neoantigens successfully identified by each of the four programs employed.*

| | ***Strelka*** | ***VarScan2*** | ***VarScan2 Unfiltered*** | ***GATK Mutect2*** | ***Lofreq*** |
|---|---|---|---|---|---|
| ***Patient 1*** | *9/9* | *7/9* | *8/9* | *9/9* | *9/9* |
| ***Patient 2*** | *4/4* | *4/4* | *4/4* | *4/4* | *4/4* |
| ***Patient 3*** | *12/13* | *12/13* | *12/13* | *12/13* | *12/13* |
| ***Patient 12*** | *1/4* | *1/4* | *1/4* | *1/4* | *1/4* |
| ***Patient 16*** | *1/4* | *0/4* | *0/4* | *1/4* | *1/4* |
| ***Total*** | *27/34* | *24/34* | *25/34* | *27/34* | *27/34* |

***Supplementary Table 2:*** *Summary of datasets used, materials used, and accession information*

| Dataset | Materials used | Accession |
|---|---|---|
| TESLA Consortium, Wells *et al*. 2020 | Raw RNAseq and WES data | https://www.synapse.org/#!Synapse:syn21048999/wiki/603788 |
| | Lists of validated neoantigens and the T cell response | Supplementary Table S4 |
| Cohen *et al*. 2015 | Raw RNAseq data | https://trace.ddbj.nig.ac.jp/DRASearch/study?acc=SRP062169 |
| | Lists of validated neoantigens and the T cell response | Supplementary Table 2 |
| Strønen *et al*. 2016 | Lists of validated neoantigens and the T cell response | Supplementary Table S8 |
| Carreno *et al*. 2015 | Lists of validated neoantigens and the T cell response | Supplementary Tables S1-S3 |
| Ott *et al*. 2018 | Lists of validated neoantigens and the T cell response | Supplementary Table 4 |
| Van Allen Dataset, Van Alen *et al*. 2015 | Raw RNAseq and WES data | Dbgap accession number (phs000452.v3.p1)) https://www.ncbi.nlm.nih.gov/projects/gap/cgi-bin/study.cgi?study_id=phs000452.v3.p1 |
| Liu Dataset, Liu *et al*. 2020 | Raw RNAseq and WES data | |

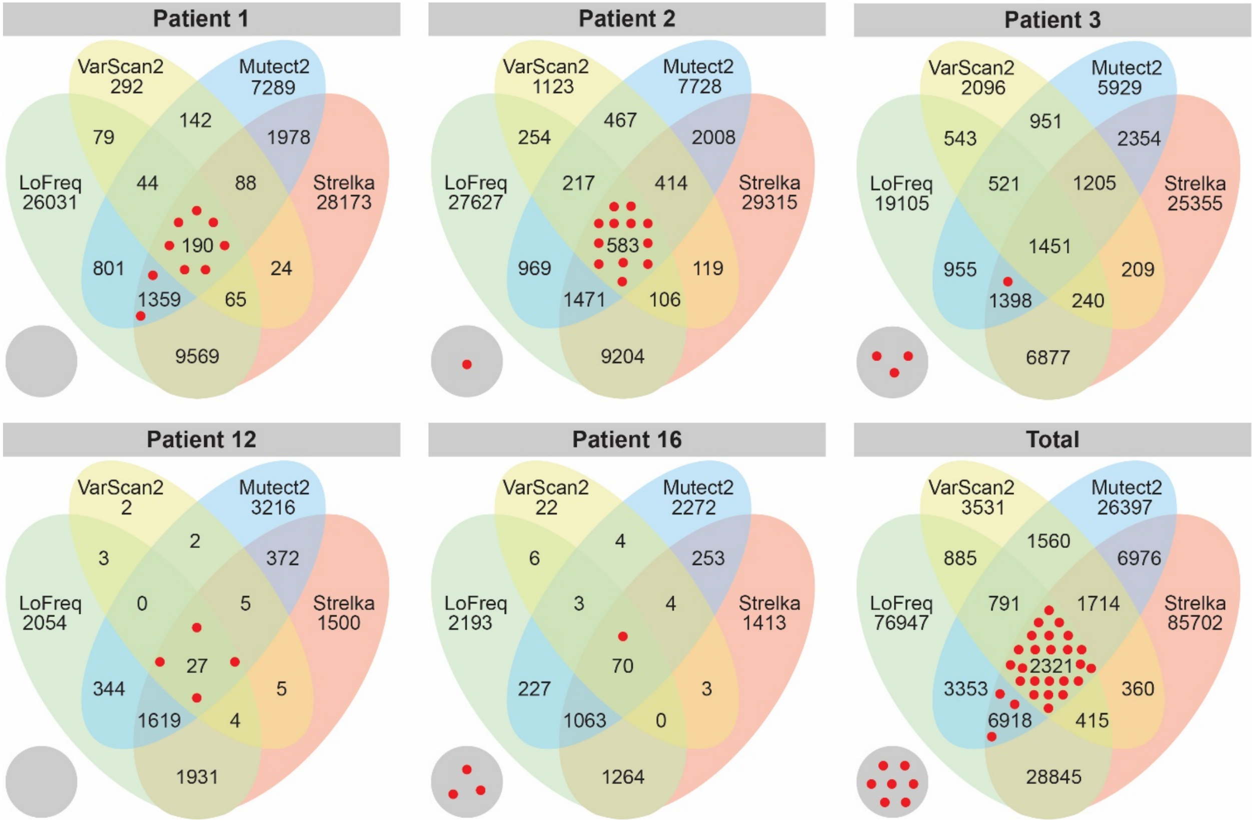


***Supplementary Figure 1. No Additional Immunogenic Mutations Identified by LoFreq.*** *Comparison of the number of neoantigens derived from single nucleotide variants (SNVs) and small insertions and deletions (indels) identified by each of four different programs (Varscan, GATK Mutect2, Strelka, and LoFreq). Red dots represent the validated immunogenic neoantigens from each patient, and the grey circle represents validated immunogenic neoantigens that were not identified by these four programs.*

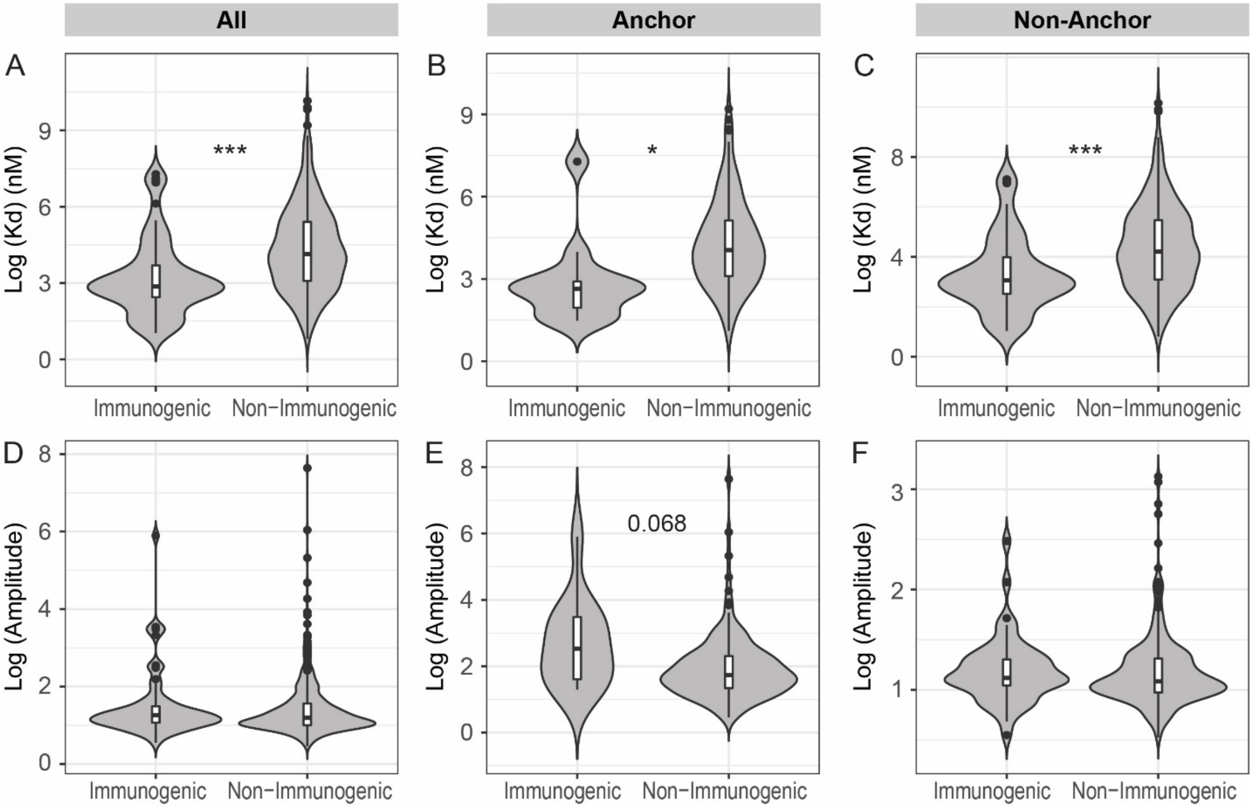


***Supplementary Figure 2. Dissociation Constant is Significantly Different for Neoantigens Derived from Mutations in Anchor and Non-anchor Residues.*** *Comparison of the dissociation constant and amplitude characteristics between immunogenic and non-immunogenic neoantigens from the TESLA consortium, Carreno, Ott, and Strønen datasets. Dissociation constant (Kd) of the MHC class I:neoantigen interaction in (A) all neoantigens, (B) neoantigens mutated at an anchor residue, and (C) neoantigens mutated at a non-anchor residue. Amplitude, defined as the ratio of MHC class I dissociation constant for the closest matched normal human peptide to the MHC class I:neoantigen dissociation constant, for (D) all neoantigens. (E) neoantigens mutated at an anchor residue, and (F) neoantigens mutated at a non-anchor residue.* $*$*:*$p<0.05$*,* $***$*:*$p<10^{-5}$*.*

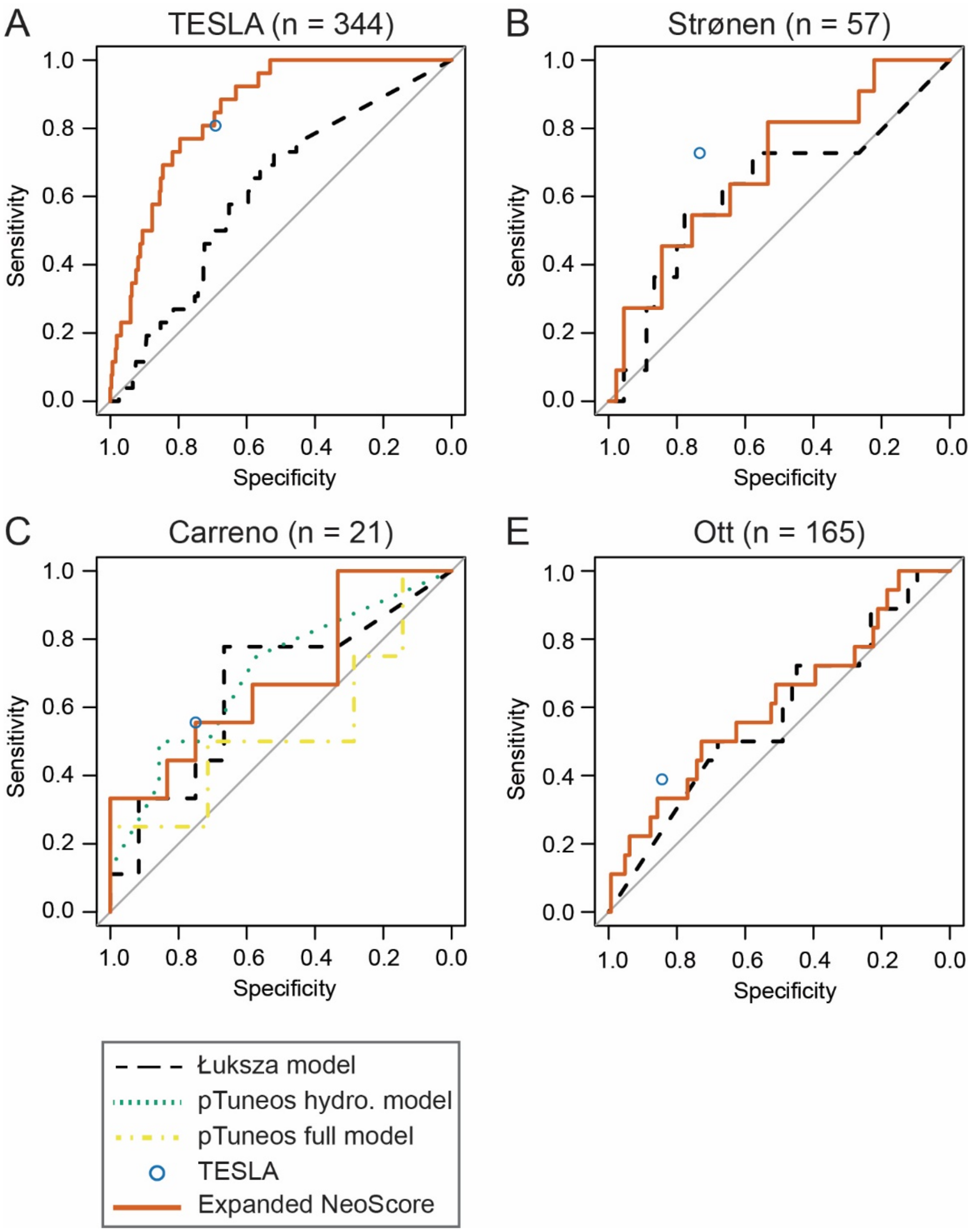


***Supplementary Figure 3. Model with All Neoantigen Characteristics Does Not Have Improved Performance.*** *Model performance when including all 9 neoantigen characteristics in expanded NeoScore. (A) Performance of the expanded NeoScore in the TESLA consortium dataset (the dataset in which the model was fit). AUC is 0.852 for the expanded NeoScore compared to 0.846 for the NeoScore. Performance of the expanded NeoScore in the (B) Strønen dataset (AUC is 0.685 for expanded NeoScore vs. 0.739 for the NeoScore), (C) Carreno dataset (AUC is 0.685 for expanded NeoScore vs. 0.739 for NeoScore), D) Ott dataset (AUC is 0.609 for expanded NeoScore vs. 0.609 for NeoScore)*
